# Extended SEIQR type model for COVID-19 epidemic and data analysis

**DOI:** 10.1101/2020.08.10.20171439

**Authors:** Swarnali Sharma, Vitaly Volpert, Malay Banerjee

## Abstract

An extended SEIQR type model is considered in order to model the COVID-19 epidemic. It contains the classes of susceptible individuals, exposed, infected symptomatic and asymptomatic, quarantined, hospitalized and recovered. The basic reproduction number and the final size of epidemic are determined. The model is used to fit available data for some European countries. A more detailed model with two different subclasses of susceptible individuals is introduced in order to study the influence of social interaction on the disease progression. The coefficient of social interaction *K* characterizes the level of social contacts in comparison with complete lockdown (*K* = 0) and the absence of lockdown (*K* = 1). The fitting of data shows that the actual level of this coefficient in some European countries is about 0.1, characterizing a slow disease progression. A slight increase of this value in the autumn can lead to a strong epidemic burst.

## 1 Introduction

The Coronavirus disease 2019 (COVID-19) pandemic is now considered as the biggest global threat worldwide. This disease is caused by severe acute respiratory syndrome coronavirus 2 (SARS-COV- 2) which belogs to a group of RNA virus causing respiratory track infection that can range mild to lathel [1]. The first outbreak of COVID-19 was noticed in Hubei province, Wuhan, China [2] in December 2019. Then it spread all over the world so rapidly that the World Health Organization (WHO) revealed the COVID-19 to be a public health emergency and identified it as a pandemic on March 11, 2020. Since December 2019, the first COVID-19 infected person was diagnosed, the COVID-19 quickly spread to all Chinese province and, as of 1 April 2020, to 200 countries and regions with the number of reported infected cases and the number of documented death reached 19 million and 717,792, respectively [3]. Among these countries, the more serious epidemic situations are in the United States (5,032,278 total cases, 162,804 death cases), Brazil (2,917,562 total cases, 98,644 death cases), India (2,030,001 total cases, 41,673 death cases), Russia (871,894 total cases, 14,606 death cases), Italy (249,204 total cases, 35,187 death cases), UK (308,134 total cases, 46,413 death cases), Spain (354,530 total cases, 28,500 death cases), Germany (215,210 total cases, 9,252 death cases), France (195,633 total cases, 30,312 death cases), and Iran (320,117 total cases, 17,976 death cases). In particular, the number of infection cases in the United States has grown very fast, the number of reported infected cases increases from 15 to 288,721 spending 82 days [3].

The most alarming part of this disease is that the symptoms of this disease are not specific and in many cases the infected person may be asymptomatic (who can infect persons without showing any symptoms of the disease). The majority of the cases initially have symptoms like common cold which includes dry cough, fever, sore throat, loss of sense of smell, headache, shortness of breathe etc. Moreover, the growth of this infection further proceed to acute respiratory distress syndrome which can lead to even death. It is also observed in an age-stratified analysis [4] that the large number of severe cases in particular for the age groups above 60 and having other medical issues like diabetics, kidney problems etc. The COVID-19 virus spreads at large extend between people when they come in close contact with each other and the virus is transmitted through expelled droplets which enter a person’s body through contact routes such as the mouth, eyes or nose. Contact with various surface is another means for contracting the virus. In the absence of a definite treatment modality like medicine or vaccine, physical distancing, wearing masks, washing hands etc. have been accepted globally as the most efficient strategies for reducing the severity and spread of this virus and gaining control over it at some extend [5]. So, the government of most of the countries had decided to go for complete lockdown from mid or end of March, 2020. This complete lockdown also affected the economy of those countries in a large extend. So, from the mid of the month of May, 2020, all the countries have taken some policies for unlocking gradually.

Mathematical models of infectious disease dynamics nowadays became a very useful and important tool for the analysis of dynamics of infectious disease, to predict the future course of an outbreak and to evaluate strategies to control an epidemic in recent years. The global problem of the outbreak of COVID-19 has attracted the interest of researchers of different areas. Mathematical modeling based on system of differential equations may provide a comprehensive mechanism for the dynamics of COVID-19 transmission. Several modeling studies have already been performed for the COVID-19 outbreak [6–11]. In [12], Lin et. al. suggested a conceptual model for the coronavirus disease 2019, which effectively catches the time line of the COVID-19 outbreak. A mathematical model for reproducing the stage-based transmissibility of a novel coronavirus by Chen et.al in [13]. Wu et al. developed a susceptible exposed infectious recovered model (SEIR) based on the reported data from December. 31st 2019 till January 28th 2020, to clarify the transmission dynamics and projected national and global spread of disease [14]. They also calculated the basic reproduction number as around 2.68 for COVID-19. Tang et al proposed a compartmental deterministic model that would combine the clinical development of the disease, the epidemiological status of the patient and the measures for intervention. Researchers also found that the amount of control reproduction number may be as high as 6.47, and that methods of intervention including intensive touch tracing followed by quarantine and isolation would effectively minimize COVID cases [15]. For the basic reproductive number 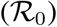, Read et al. reported a value of 3.1 based on the data fitting of an SEIR model, using an assumption of Poisson-distributed daily time increments [16]. S. Zhao et al. estimated the mean 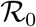 for 2019-nCoV in the early phase of the outbreak ranging from 3.3 to 5.5 (likely to be below 5 but above 3 with rising report rate) [17], which appeared slightly higher than those of SARS-CoV 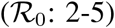 [18]. A report by Cambridge University has indicated that India’s countrywide three-week lockdown would not be adequate to prevent a resurgence of the new coronavirus epidemic that could bounce back in months and cause thousands of infections [19]. They suggested that two or three lockdowns can extend the slowdown longer with five-day breaks in between or a single 49-day lock-down. Data-driven mathematical modeling plays a key role in disease prevention, planning for future outbreaks and determining the effectiveness of control. Several data-driven modeling experiments have been performed in various regions [15]. In [20], a compartmental mathematical model to try to understand the outbreak of COVID-19 in Mexico. This data driven analysis would let us compare how different the outbreak will be in the two studied regions. By this approach, authorities can plan a health care program and control the spread even with limited resources. In Rojas et al. [21], the authors estimated the value of 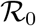 which helped them to predict that in the city of Cali the outbreak under current intervention of isolation and quarantine will last for 5-6 months and will need around 3500 beds on a given during the peak of the outbreak. Some other relevant works where the basic reproduction number is estimated for different countries can be found in [22–28].

The most common mathematical formulations which represent the individual transition in a community between ‘compartments’ describe the situation of individual infection with a significant insight. These models of compartmental disease segregate a population into groups depending on each individual’s infectious state and related population sizes with respect to time. There is a wide range of mathematical models and approaches adopted by different researchers to develop viable mathematical model to understand the propagation of disease spread for COVID-19 with different model assumptions. In our present work we have proposed and analyzed an ordinary differential equation model to study the COVID-19 disease propagation which consists with initially six compartments namely susceptible, exposed, infected, quarantined, hospitalised and recovered. We have divided the exposed compartment into two sub-compartments *E*_1_ and *E_2_* depending on their infectiousness and also divided the infected class into two sub-compartments namely asymptomatic (*I_a_*) and symptomatic (*I_s_*). Interesting and significant contribution of our work is the consideration of time dependent rate of infection over various periods of time. This variation is adopted into the model in order to capture the effect of lockdown, social distancing etc. which plays a crucial role to reduce the disease spread. Next we have discussed the basic properties of our model and calculated the basic and controlled reproduction number. Final size of the epidemic is also described. Next we have divided susceptible, exposed and infected classes into two sub-classes each based on their classification or behaviour which is directly responsible for the alteration of rate of disease spread. The classification or division into two different groups may be due to the different age groups, different implementation of distancing measures, proper and improper use of face mask and so on. Then we have calculated the reproduction number and maximum size of the epidemic for the new model. Next we have copulated the sensitivity index to identify the parameters of greater interest and then fitted those parameter values with the data of total cumulative cases of COVID-19 for different countries (Germany, Italy, Spain, and UK). Then we have shown that with those best fitted parameter values our model simulation is matching well with the 95% confidence interval of the daywise cumulative and daily case data which proved the viability of our model system. This also depict the fact that nature of the disease transmission is different for different countries depending on the protective measures and policies taken by government, different age distribution of the population, lack of consciousness etc.

## 2 Mathematical Model – 1

In the following, we consider a dynamic SEIQR type model for the COVID-19 disease propagation. Basically the model consists with Susceptible (*S*), Exposed (*E*), Infected (*I*), Quarantined (*Q*), hospitalized (*J*) and Recovered (*R*) class. In the context of COVID-19, the exposed class is divided into two subclasses namely non-infectious (*E*_1_) and infectious (*E*_2_) and the infected class is divided into two sub- classes namely asymptomatic (*I_a_*) and symptomatic (*I_s_*), where *N* = *S* + *E*_1_ + *E*_2_ + *I_a_* + *I_s_* + *Q* + *J* + *R* and *N* is not fixed since deceased individuals are not considered in the model. The model assumptions are given as follows:

**Susceptible population** *S*(*t*): This subpopulation will decrease after an infection due to the interaction with an symptomatic infected individual (*I_s_*), asymptomatic infected individual (*I_a_*), infectious exposed individual (*E*_2_), quarantine (*Q*) or hospitalised one (*J*). The transmission coefficients will be *βI_s_*, *βp*_1_*I_a_*, *βp*_2_*E*_2_, *βp*_3_*Q* and *βp*_4_*J* respectively. Here *β* is rate of infection per unit of time by the symptomatic infected, *p*_1_, *p*_2_, *p*_3_ and *p*_4_ are the reduction factor of infectivity by *I_a_*, *E*_2_, *Q* and *J* respectively compared to *I_s_* and satisfy the restriction 0 ≤ *p_j_ <* 1, *j* = 1, 2, 3,4. The rate of change of the susceptible population is expressed in the following equation:

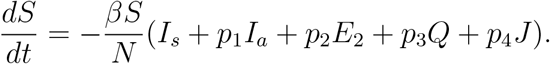

**Exposed polulation** *E*(*t*): The exposed population (in the incubation period) is divided into two sub classes: (i) exposed population who are at the beginning of the incubation period and cannot spread the disease (*E*_1_(*t*)) and (ii) exposed population who are at the end of the incubation period and can spread the disease (*E*_2_(*t*)). The transfer mechanism from the class *S*(*t*) to the class *E*_1_(*t*) is guided by the function 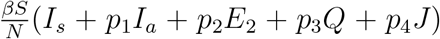 and from the class *E*_1_(*t*) to the class *E*_2_(*t*) is guided by *μ*E_1_, where *μ* is the rate at which individuals of *E*_1_ class become infectious exposed (*E*_2_). The population *E*_2_ will decrease due to the transfer into the infected population with rate *δ*. Thus the rate of change of the exposed population is expressed in the following two equations:

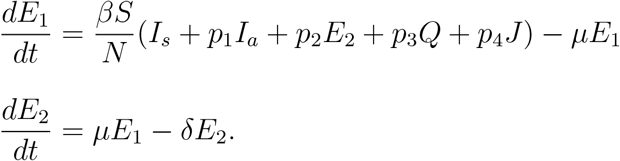

**Infected polulation** *I*(*t*): The infected population is divided into two subclasses (i) asymptomatic (having no symptoms) (*I_a_*) and (ii) symptomatic (having symptoms) (*I_s_*). The transfer mechanism from the class *E*_2_(*t*) to the class *I_a_*(*t*) is guided by the function (1 − *σ*) *δE*_2_ and from the class *E*_2_(*t*) to the class *I_s_*(*t*) is guided by the function *σδ*E_2_ where *σ* (0 < *σ* < 1) is the fraction of *E*_2_ that becomes symptomatic infected (*I_s_*). The population *I_a_* will decrease due to the transfer into the recovered population with a rate *η* and *I_s_* will decrease with a rate (*ρ*_1_ + *ζ*_1_ + *ζ*_2_ + *ζ*_3_), where *ρ*_1_ is the rate of mortality due to the infection, *ζ*_1_, *ζ*_2_ and *ζ*_3_ are the rate at which *I_s_* becomes quarantined, recovered and hospitalised respectively. Thus the rate of change of the infected population is expressed in the following two equations:

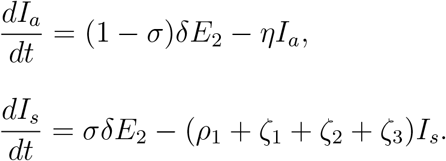

**Quarantined population** *Q*(*t*): This subpopulation will increase due to the transfer from the class *I_s_*(*t*) with a rate *ζ*_1_ and will decrease with a rate (ξ_1_ + ξ_2_), where ξ_1_ and ξ_2_ are the rates at which *Q* becomes hospitalized and recovered respectively. Thus the rate of change of the quarantined population is expressed in the following equation:

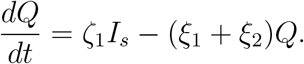

**Hospitalized population** *J*(*t*): This subpopulation will increase due to the transfer from the classes *I_s_*(*t*) and *Q*(*t*) with rates *ζ*_3_ and ξ_1_ respectively and will decrease with a rate (*ρ*_2_ + *ν*), where *ρ*_2_ is the rate of mortality due to the infection and *ν* is the rate at which *J* becomes recovered. Thus the rate of change of the quarantined population is expressed in the following equation:

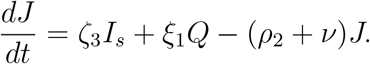

**Recovered population** *R*(*t*): This subpopulation will increase due to recovery from the disease from the classes *I_a_*(*t*), *I_s_*(*t*), *Q*(*t*) and *J*(*t*) with rates *η*, *ζ*_2_, ξ_2_ and *ν* respectively. Thus the rate of change of the recovered population is expressed in the following equation:

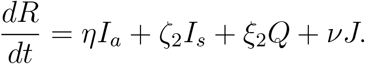

Hence, the system of differential equations that will model the dynamics of coronavirus spread is:

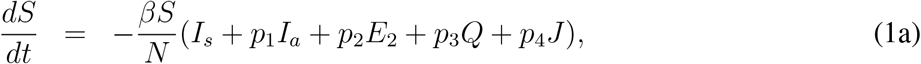

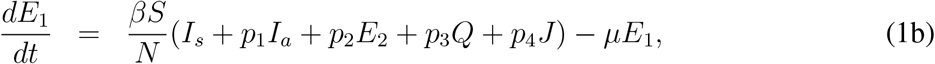

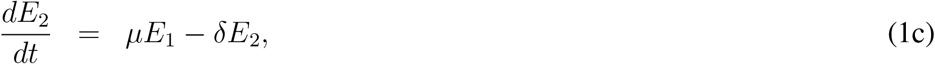

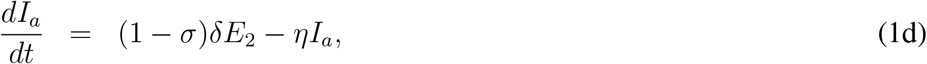

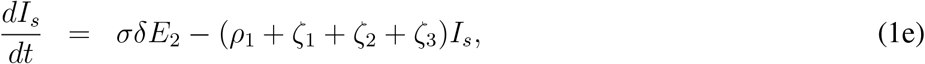

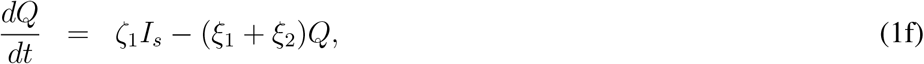

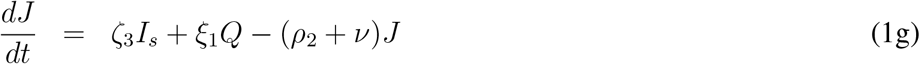

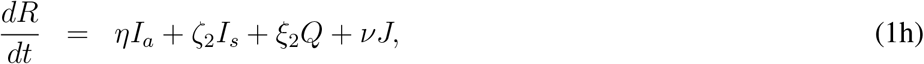

subjected to non-negative initial conditions *S*(0); *E*_1_(0); *E*_2_(0); *I_a_*(0); *I_s_*(0);*Q*(0); *J*(0);*R*(0) ≥ 0.

Interpretation for the parameters involved with the model (1) is summarized in Table 1 for a quick reference. A schematic diagram for the transmission of disease and progression of the individuals from one compartment to another is provided in Fig. 3.

**Figure 1:**
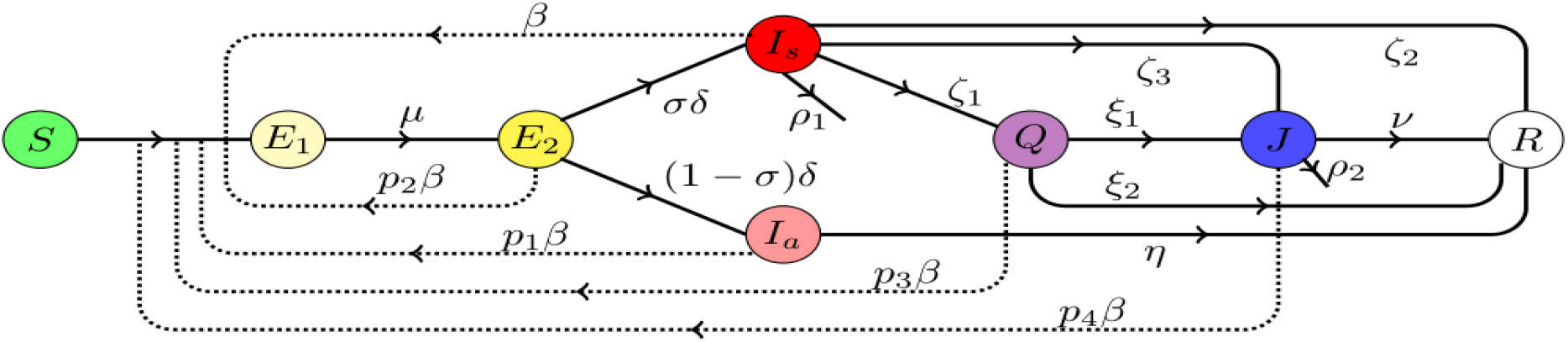
Schematic diagram for the progression of disease described by the model (1). Solid arrows represent the transfer from one compartment to another while the dotted line with arrow denote the compartments responsible for disease transmission. Associated rates are mentioned accordingly.

We have written the basic model with *β* as constant for the simplicity of forthcoming mathematical calculations. However, for numerical simulations and in order to fit the numerical results with available data [3], we will consider *β* = *β*(*t*) as a function of time in order to model the effect of lockdown. In reality the rate of infection is not a constant throughout the epidemic rather it changes time to time due to variable social behavior. Although it is difficult to obtain actual pattern of variation with respect to time but we have considered this variation in order to model the lowering in rate to infection due to the lowering of social contancts through lockdown.

**Table 1:**
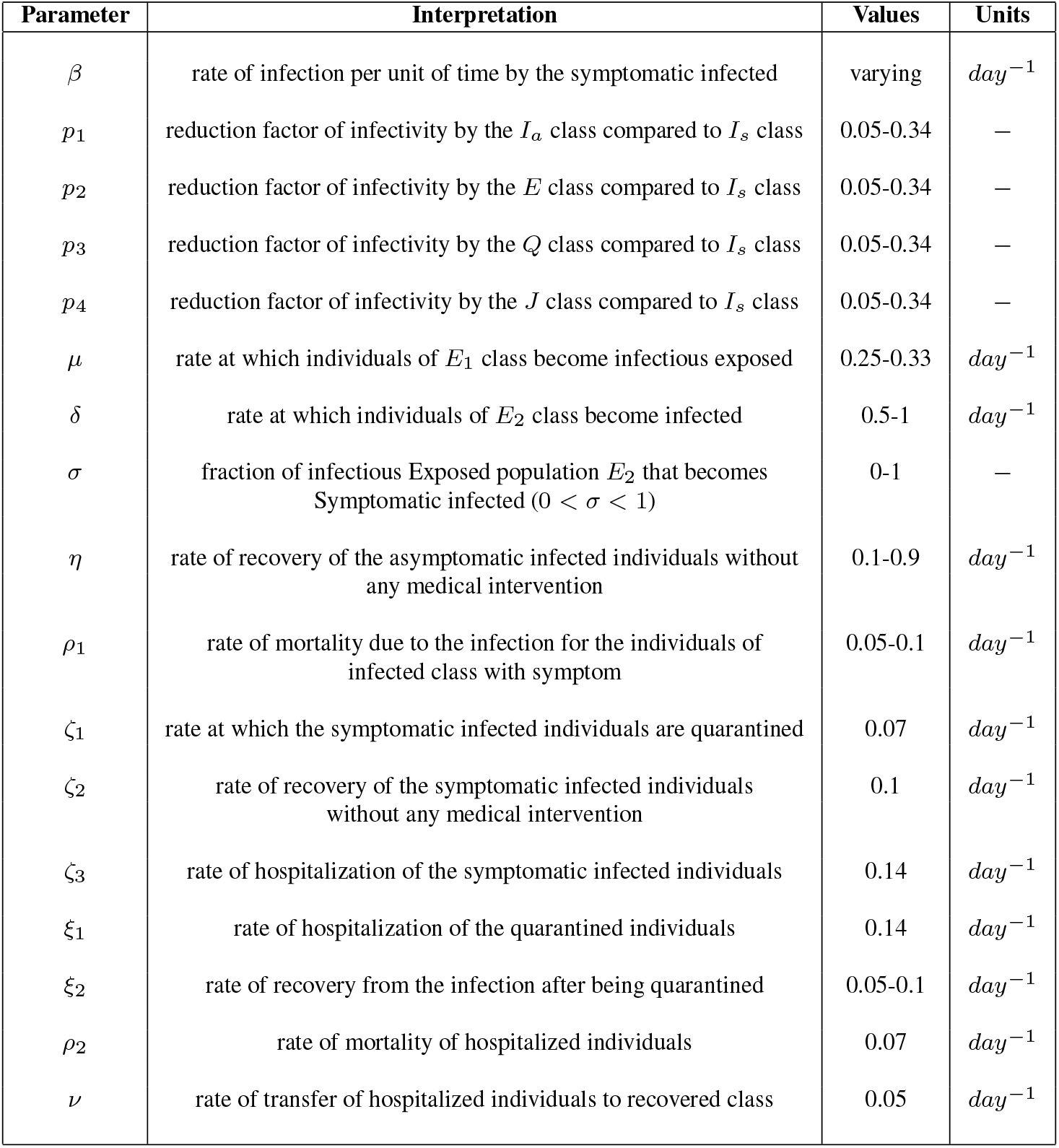
Description of parameters involved with the model (1). Range of parameter values are obatined from the references [21,25,26,30-32].

| Parameter | Interpretation | Values | Units |
| --- | --- | --- | --- |
| $\beta$ | rate of infection per unit of time by the symptomatic infected | varying | $day^{-1}$ |
| $p_1$ | reduction factor of infectivity by the $I_a$ class compared to $I_s$ class | 0.05-0.34 | — |
| $p_2$ | reduction factor of infectivity by the $E$ class compared to $I_s$ class | 0.05-0.34 | — |
| $p_3$ | reduction factor of infectivity by the $Q$ class compared to $I_s$ class | 0.05-0.34 | — |
| $p_4$ | reduction factor of infectivity by the $J$ class compared to $I_s$ class | 0.05-0.34 | — |
| $\mu$ | rate at which individuals of $E_1$ class become infectious exposed | 0.25-0.33 | $day^{-1}$ |
| $\delta$ | rate at which individuals of $E_2$ class become infected | 0.5-1 | $day^{-1}$ |
| $\sigma$ | fraction of infectious Exposed population $E_2$ that becomes Symptomatic infected ($0 < \sigma < 1$) | 0-1 | — |
| $\eta$ | rate of recovery of the asymptomatic infected individuals without any medical intervention | 0.1-0.9 | $day^{-1}$ |
| $\rho_1$ | rate of mortality due to the infection for the individuals of infected class with symptom | 0.05-0.1 | $day^{-1}$ |
| $\zeta_1$ | rate at which the symptomatic infected individuals are quarantined | 0.07 | $day^{-1}$ |
| $\zeta_2$ | rate of recovery of the symptomatic infected individuals without any medical intervention | 0.1 | $day^{-1}$ |
| $\zeta_3$ | rate of hospitalization of the symptomatic infected individuals | 0.14 | $day^{-1}$ |
| $\xi_1$ | rate of hospitalization of the quarantined individuals | 0.14 | $day^{-1}$ |
| $\xi_2$ | rate of recovery from the infection after being quarantined | 0.05-0.1 | $day^{-1}$ |
| $\rho_2$ | rate of mortality of hospitalized individuals | 0.07 | $day^{-1}$ |
| $\nu$ | rate of transfer of hospitalized individuals to recovered class | 0.05 | $day^{-1}$ |

### 2.1 Positivity and boundedness

A viable mathematical model for epidemiology must ensure that the solutions of the model under consideration remain non-negative once started from an interior point of the positive cone and remains bounded at all future time. The model considered here is not a completely new and several close versions are available in various articles on mathematical epidemiology. However, for the completeness we just state the relevant result and a brief outline for the proof is provided at the appendix. For this purpose we consider that the model (1) is subjected to the initial conditions (*S*(0), *E*_1_(0), *E*_2_(0), *I_a_*(0), *I_s_*(0), *Q*(0), *J*(0), 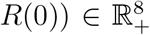, where positive cone of 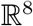 is 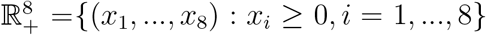. First we prove that the solution of the system (1) remains within 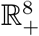 at all future time once started from a point within 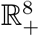 following the approach outlined in [29].

#### Theorem 1.

*The system (1) is invariant in* 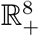.

Proof.

See appendix.

In order to close the model (1), we can introduce the compartment for number of COVID related deaths as follows

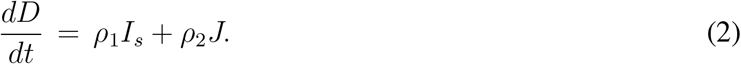

Now if we write *N_D_* = *S* + *E*_1_ + *E*_2_ + *I_a_* + *I_s_* + *Q* + *J* + *R* + *D*, then from (1) and (2), we find

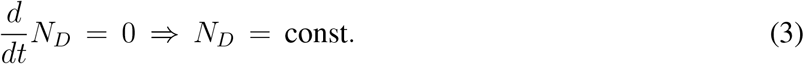

With the previous urgument we can prove *D*(*t*) ≥ 0 and hence 0 < *N* = *N_D_* implies *N* is bounded above, consequently all the constituent variables are also bounded.

### 2.2 Basic and controlled reproduction numbers

In this section, first we find the basic reproduction number for the model (1) in the absence of any control measure like isolation, quarantine and hospitalization. For this purpose we assume that the model consists with Susceptible, Exposed, Infected and Recovered class only. The Quarantined and Hospitalized classes are absent. Based upon this assumption we can find the following reduced model:

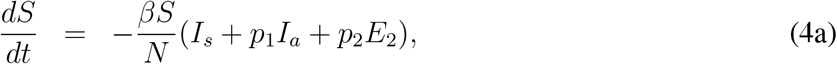

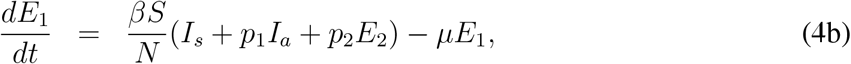

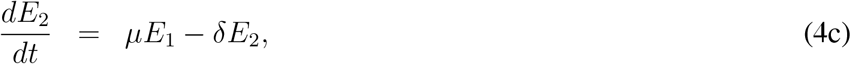

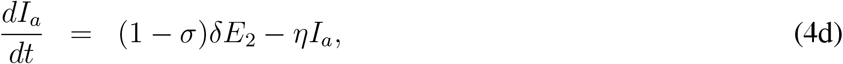

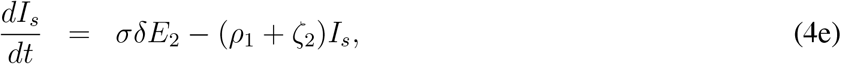

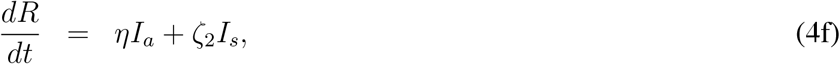

subjected to non-negative initial conditions *S*(0), *E*_1_(0), *E*_2_(0), *I_a_*(0), *I_s_*(0), *R*(0) ≥ 0. For this model *N* = *S* + *E*_1_ + *E*_2_ + *I_a_* + *I_s_*+ *R*. Here we calculate the basic reproduction number for above model following the next generation matrix approached as introduced in [33]. The disease free equilibrium point is given by (*N*, 0, 0, 0, 0, 0), the derivation of basic reproduction number is based upon the stability condition of disease free equilibrium point for the model (4). First we rearrange the compartments of the model (4) such that first *m*(= 4) equations are related to infected compartments from epidemiological point of view. Susceptible and recovered compartments will be placed at the end. From (4), we can write

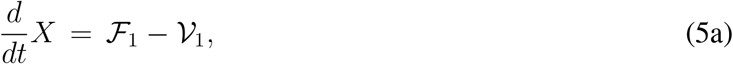

where *X* = [*E*_1_;*E*_2_; *I_a_*; *I_s_*; *S*; *R*]*^T^* ≡ [*x*_1_; *x*_2_; *x*_3_; *x*_4_; *x*_5_; *x*_6_]*^T^* and 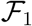, 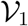 are defined by

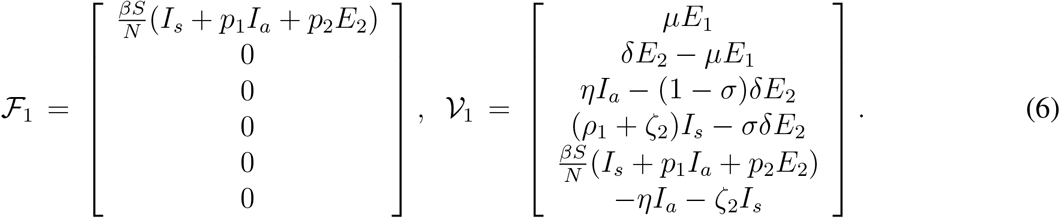

Here the matrix 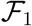 consists of the terms involved with the appearence of new infection at all the compartments and 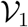 contains the terms representing entry and exit of all other individuals, rather than direct infection, at all compartments. For the calculation of basic reproduction number now we need to evaluate two matrices *F*_1_ and *V*_1_, which are defined by

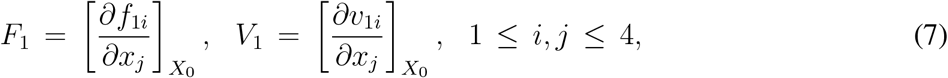

where *X*_0_ = (0; 0; 0; 0; *N*; 0)*^T^*. Here we should mention that 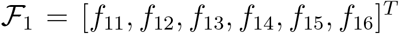 and 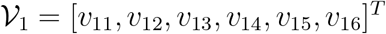 to avoid any confusion. Hence we can calculate

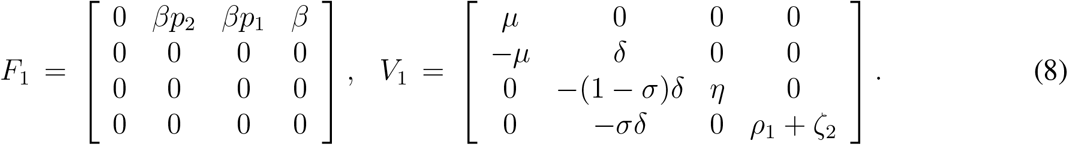

The largest eigenvalue of the next generation matrix *F*_1_*V*_1_^−1^ model (4), (see [33,34]). Now, we can calculate,

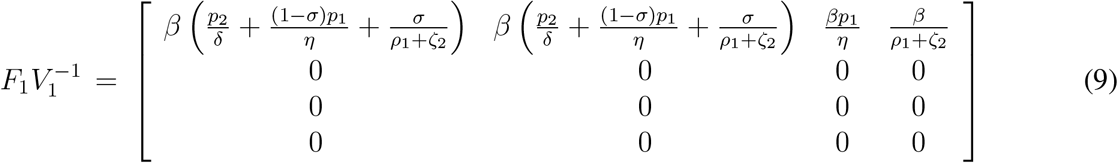

The basic reproduction number for the model (4) is denoted by 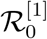 and is given by

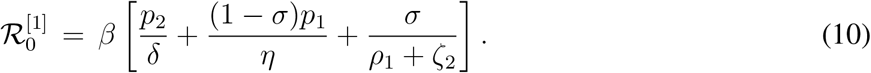

Here the superscript ‘[1]’ stands for the first model considered in this manuscript, that is for the model (1). This basic reproduction number is the sum of three terms, which represents the number of secondary infections produced by an infectious exposed individual, an asymptomatic infected individual, and a symptomatic infected individual respectively. 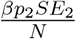 is the incidence of an exposed individuals who are at the end of the incubation period and can spread the disease. The number of secondary infection produced by an individual of *E*_2_ compartment in an entirely susceptible population is *βp*_2_ per unit of time. An individual spents an average 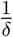 units of time in *E*_2_ compartment. Hence the number of secondary infection produced by an individual of *E*_2_ compartment is 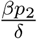. The number of secondary infections produced by the individuals of *I_a_* and *I_s_* compartments in an entirely susceptible population, per unit of time, are *β* (1 − *σ*)*p*_1_ and *β σ* a respectively. The average time units spend by the asymptomatic and symptomatic infectious individuals with their respective compartments are 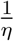 and 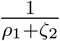. Hence the number of secondary infection produced by the individuals of *I_a_* and *I_s_* compartments are 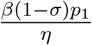 and 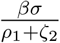 respectively.

Henceforth we follow the similar notation and approach to calculate other relevant reproduction numbers without providing much description. Now we calculate the controlled reproduction number for the model (1).

The model (1) can be written as follows

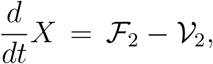

*X* = [*E*_1_, *E*_2_, *I_a_*, *I_s_*, *Q*, *J*, *S*, R]*^T^* ≡ [*x*_1_, *x*_2_, *x*_3_, *x*_4_, *x*_5_, *x*_6_, *x*_7_, *x*_8_]*^T^* with six (*m* = 6) compartments contribute to the propagation of infection. 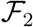 and 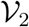 are given by

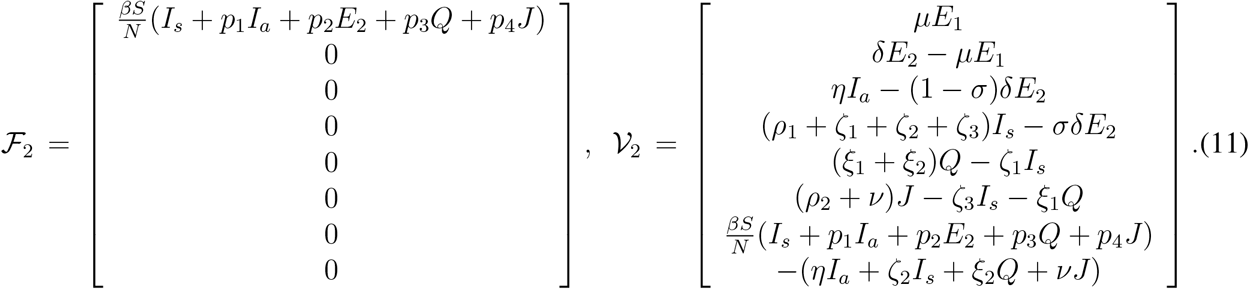

We can define following two matrices

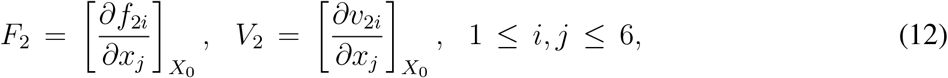

where *X*_0_ = (0,0,0,0,0,0, *N*, 0). These two matrices are given by

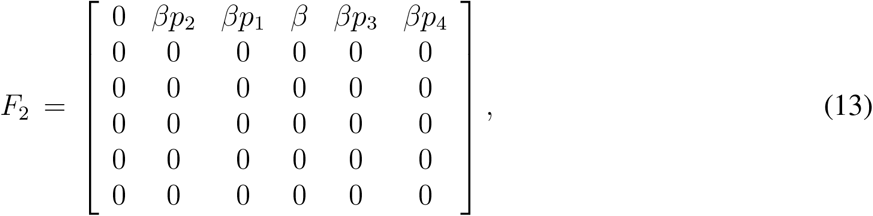

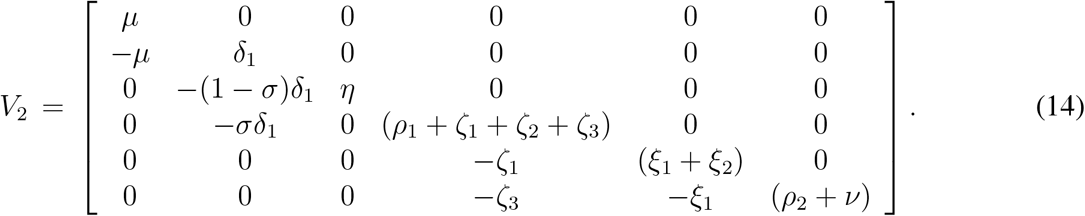

The controlled reproduction number for the model (1) is given

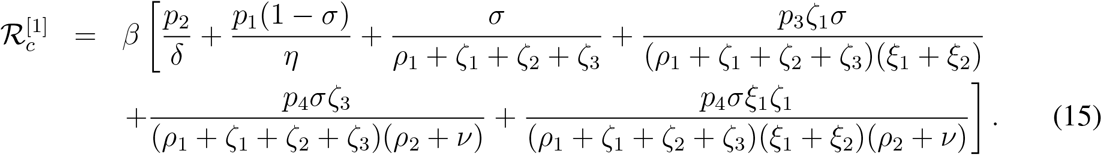

The different terms involved with 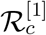 can be explained in a similar way as given above, interested readers can see [35, 36] for detailed discussion. It is important to mention here that substituting *p*_3_ = *p*_4_ = *ζ*_1_ = *ζ*_3_ = 0 we can find 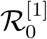 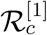.

### 2.3 Final sizes of epidemic

In order to understand the final sizes of the epidemic, we find three impotant quantites *S_f_*, *R_f_* and *D_f_*. First we calculate the final size of the susceptible compartment that is *S_f_*.

We integrate the equation for *S* (*i.e*. (1a)) between *t* = 0 to *t* = *t_f_* and find

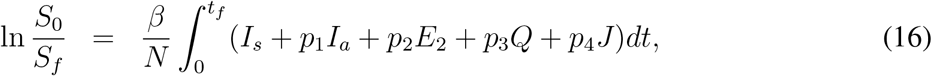

where *S*_0_ and *S_f_* denotes initial and final size of the susceptible population.

Now we assume that the model (1) is subjected to the initial condition that *S*_0_, *E*_10_ > 0 and all other components are absent at the initial time point *t* = 0. Consequently *S*_0_ + *E*_10_ = *N*. Now integrating (1c) between *t* = 0 and *t* = *t_f_* and with the assumption that *E*_20_ = *E*_2_*_f_* = 0, we find

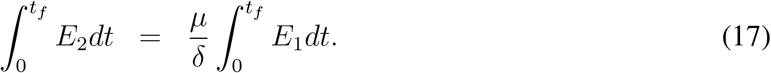

Similarly from (1d) we find the following result,

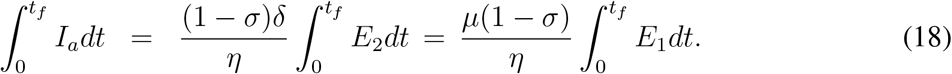

Proceeding in a similar way, from equations (1e) - (1g) and using above results we find,

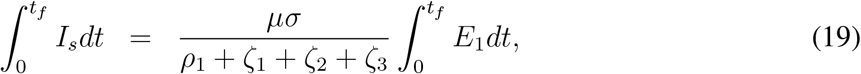

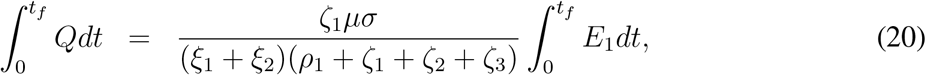

and

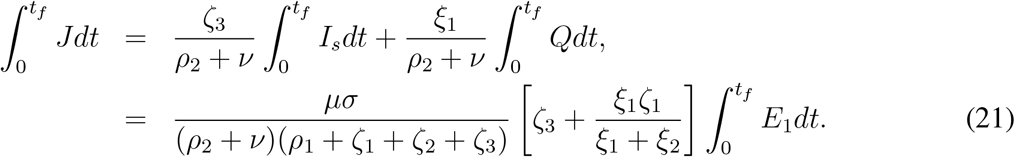

Now if we add two equations (1a) and (1b), and then integrating we find

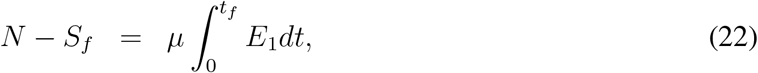

where *S*_0_ = *N* and we assume *E*_10_, *E*_1_*_f_* = 0. Now using the results (17) - (21), from (23) we find

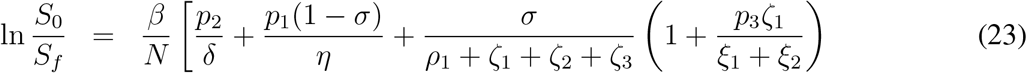

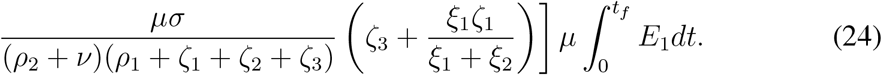

Finally using (22) and the expression for 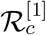, we obtain the final size of the epidemic as follows

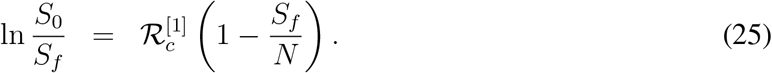

At the begining of the epidemic without any loss of generality we can assume that the entire population is susceptible and hence *N* = *S*_0_. Using 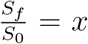 in above equation we get the following equation

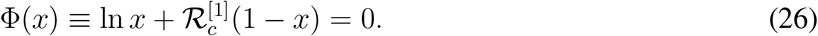

Above equation possesses a solution within the interval (0,1) when 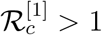 and root of the equation Φ(*x*) = 0 gives the final size of the epidemic. On the other hand the question of final size of the epidemic will not arise in case of 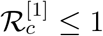 as there is disease growth. This claim can be verified from the Fig. 2.

**Figure 2:**
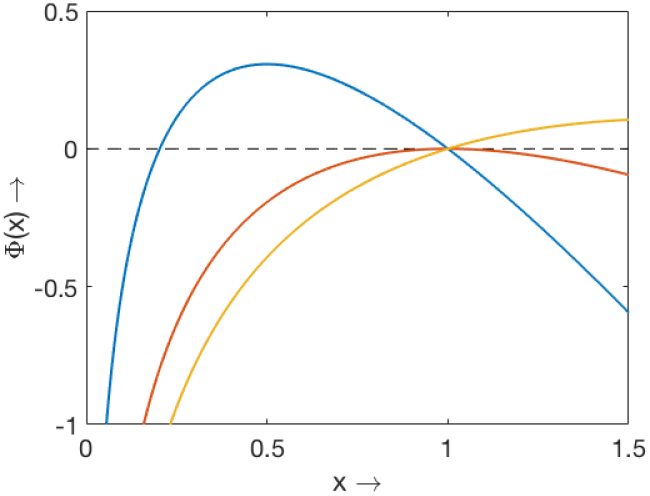
Plot of Φ(*x*) for three different values of 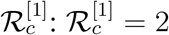 (blue), 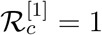 (red) and 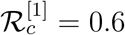 (yellow).

To understand the final size of the deceased compartment we need to calculate *D_f_* and *R_f_*. Clearly *R*_0_ = 0 and *D*_0_ = 0, hence by integrating the equations for recovered and deceased compartments between *t* = 0 to *t* = *t_f_* we find

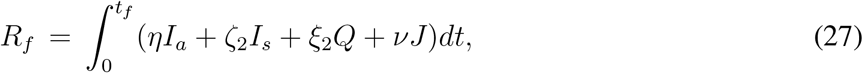

and

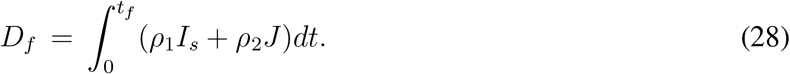

Using the results (18) – (21), we find from above two equations

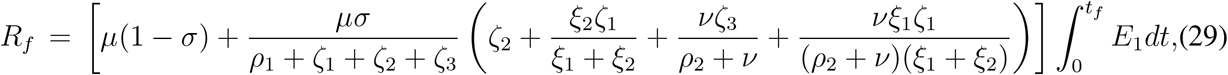

and

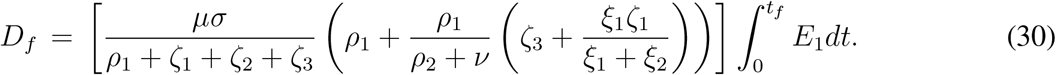

If we define

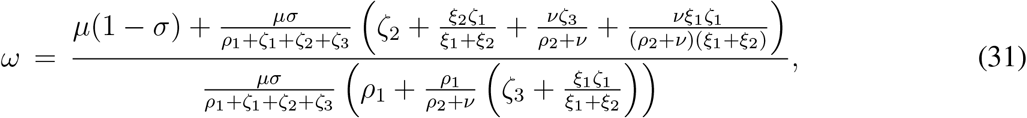

then

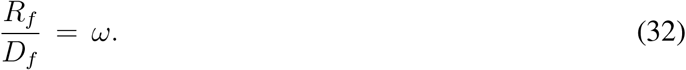

## 3 Mathematical Model - 2

In the following, we have divided the susceptible population (*S*), Exposed population (cannot spread infection (*E*_1_) and can spread infection (*E*_2_)), asymptomatic infective (*I_a_*) and symptomatic infective (*I_s_*) of model (1) into two subclasses, namely *S*_1_, *S*_2_, *E*_11_, *E*_21_, *E*_12_, *E*_22_, *I_a_*_1_, *I_a_*_2_, *I_s_*_1_, *I_s_*_2_ respectively based on their classification or behaviour which is directly responsible for the alteration of rate of disease spread. The classification or division into two different groups may be due to the different age groups, different implementation of distancing measures, proper and improper use of face mask and so on. The total population size can be written as *N* = *S*_1_ + *S*_2_ + *E*_11_ + *E*_21_ + *E*12 + *E*_22_ + *I_a_*_1_ + *I_a_*_2_ + *I_s_*_1_ + *I_s_*_2_ + *Q* + *J* + *R* and *N* is not fixed since deceased compartment is not included in the model. The assumptions for the extended model formulation are given as follows:

**Susceptible population** *S*(*t*): This subpopulation is divided into two subclasses *S*_1_ and *S*_2_. *S*_1_ will decrease after an infection due to the interaction with an symptomatic infected individual (*I_s_*_1_, *I_s_*_2_) with transmission coefficients *β*_11_*I_s_*_1_, *β*_12_*I_s_*_2_ respectively, asymptomatic infected individual (*I_a_*_1_, *I_a_*_2_) with transmission coefficients *β*_11_*p*_11_*I_a_*_1_, *β*_12_*p*_21_*I_a_*_2_ respectively, infectious exposed individual (*E*_12_, *E*_22_) with transmission coefficients *β*_11_*p*_12_*E*_12_, *β*_12_*p*_22_*E*_22_ respectively, quarantine (*β*_11_) with transmission coefficients *β_Q_Q* or hospitalised one (*J*) with transmission coefficients *β_J_J*. Here *β*_11_, *β*_12_ are rates of infection per unit of time by the symptomatic infected *I_s_*_1_, *I_s_*_2_ respectively, *β*_Q_, *β*_J_ are rates of infection per unit of time by the quarantined and hospitalized population (*Q*, *J*) respectively, *p*_11_, *p*_12_, *p*_21_ and *p*_22_ are the reduction factor of infectivity by *I_a_*_1_, *E*_12_, *I_a_*_2_ and *E*_22_ respectively compared to *I_s_*_1_ and *I_s_*_2_ and satisfy the restriction 0 ≤ *p_j_* < 1, *i*, *j* = 1, 2. *S*_2_ will decrease after an infection due to the interaction with an symptomatic infected individual (*I_s_*_1_, *I_s_*_2_) with transmission coefficients *β*_21_*I_s_*_1_, *β*_22_*I_s_*_2_ respectively, asymptomatic infected individual (*I_a_*_1_, *I_a_*_2_) with transmission coefficients *β*_21_*p*_31_*I_a_*_1_, *β*_22_*p*_41_*I_a_*_2_ respectively, infectious exposed individual (*E*_12_, *E*_22_) with transmission coefficients *β*_21_*p*_32_*E*_12_, *β*_22_*p*_42_*E*_22_ respectively, quarantine (*Q*) with transmission coefficients *β_Q_Q* or hospitalised one (*J*) with transmission coefficients *β*_J_ *J*. Here *β*_21_, *β*_22_ are rates of infection per unit of time by the symptomatic infected *I_s_*_1_, *I_s_*_2_ respectively, *β_Q_*, *β*_J_ are rates of infection per unit of time by the quarantined and hospitalized population (*Q*, *J*) respectively, *p*_31_, *p*_32_, *p*_41_ and *p*_42_ are the reduction factor of infectivity by *I_a_*_1_, *E*_12_, *I_a_*_2_ and *E*_22_ respectively compared to *I_s_*_1_ and *I_s_*_2_ and satisfy the restriction 0 ≤ *p_ij_* < 1, *i* = 3,4, *j* = 1, 2. The rate of change of the susceptible population is expressed in the following two equations:

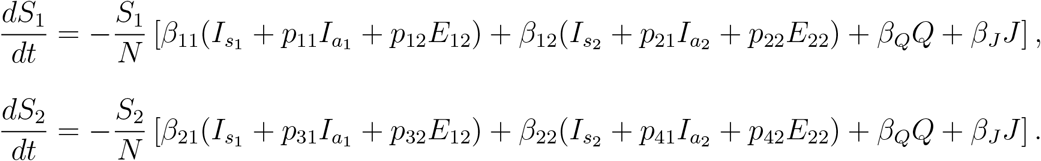

The rate of change of two groups of the exposed compartments, who are not infectious, is described by the follwoing two equations

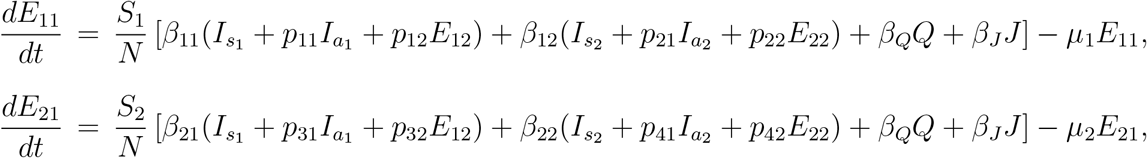

where *μ*_1_ and *μ*_2_ are the rates at which *E*_11_ and *E*_21_ progress to the infectious exposed compartments *E*_12_ and *E*_22_ respectively. The infectious exposed individuals leave the classes at rates *S*_1_ and *S*_2_ respectively and hence their rate of change with respect to time is given by

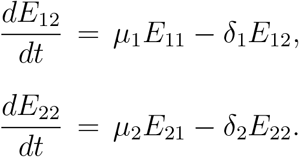

The governing equations for the two groups of asymptomatic and symptomatic infected classes are given as follows:

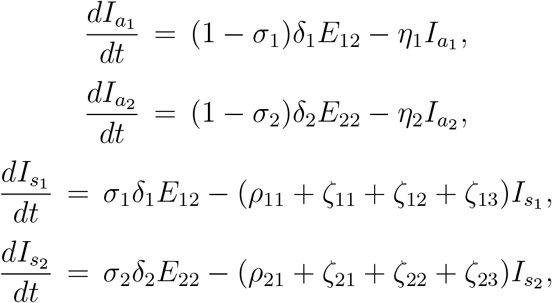

where the parameters *σ_j_*, *ρ_j_*_1_, *ζ_j_*_1_, *ζ_j_*_2_ and *ζ_j_*_3_(*j* = 1, 2) have the same interpretation as that of *σ*, *ρ*_1_ *ζ*_1_ *ζ*_2_ and *ζ*_3_ as described in Table 1 for model (1).

Distinction for quarantined, hospitalized and recovered compartments are not required as first two groups are under indirect and direct medical interventions. Finally including the governing equations for quarantined, hospitalized and recovered compartments and using previous equations we get the final two-group infection model as follows,

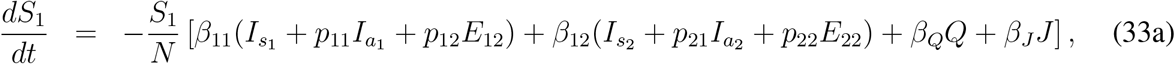

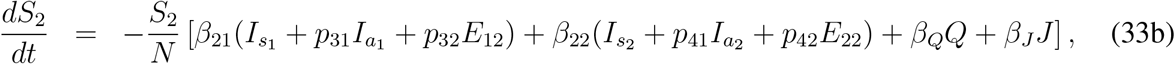

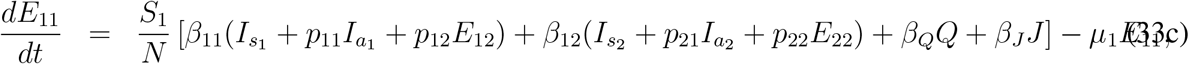

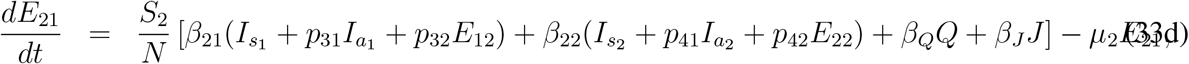

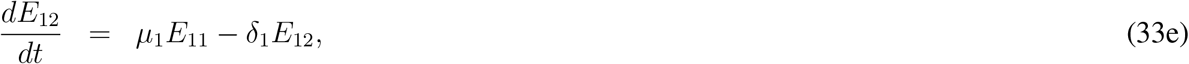

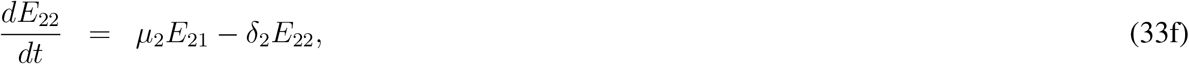

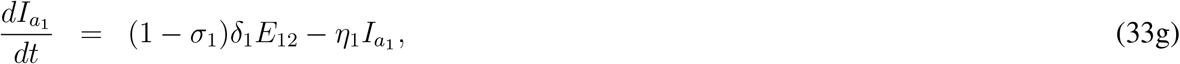

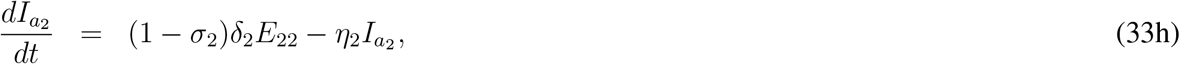

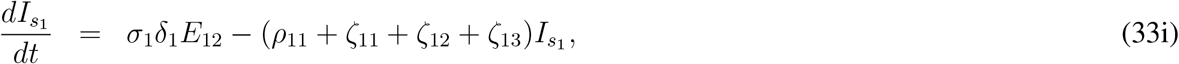

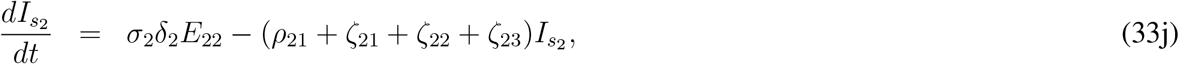

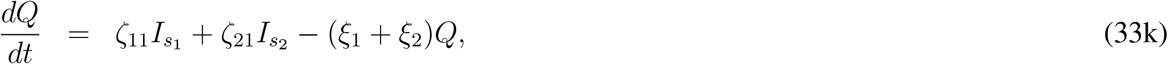

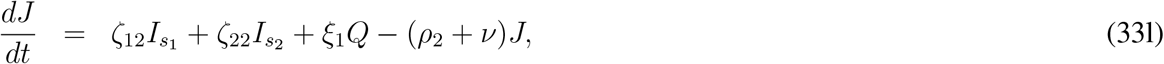

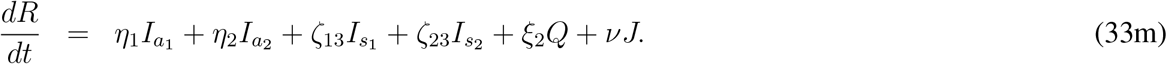

Here *ρ*_11_, *ρ*_21_, *ξ*_1_ and *ρ*_2_ are the disease related death rates for the compartments *I_s_*_1_, *I_s_*_2_, *Q* and *J* respectively. A schematic diagram is presented in Fig. 3 without the rate constants in order to avoid clumsiness

**Figure 3:**
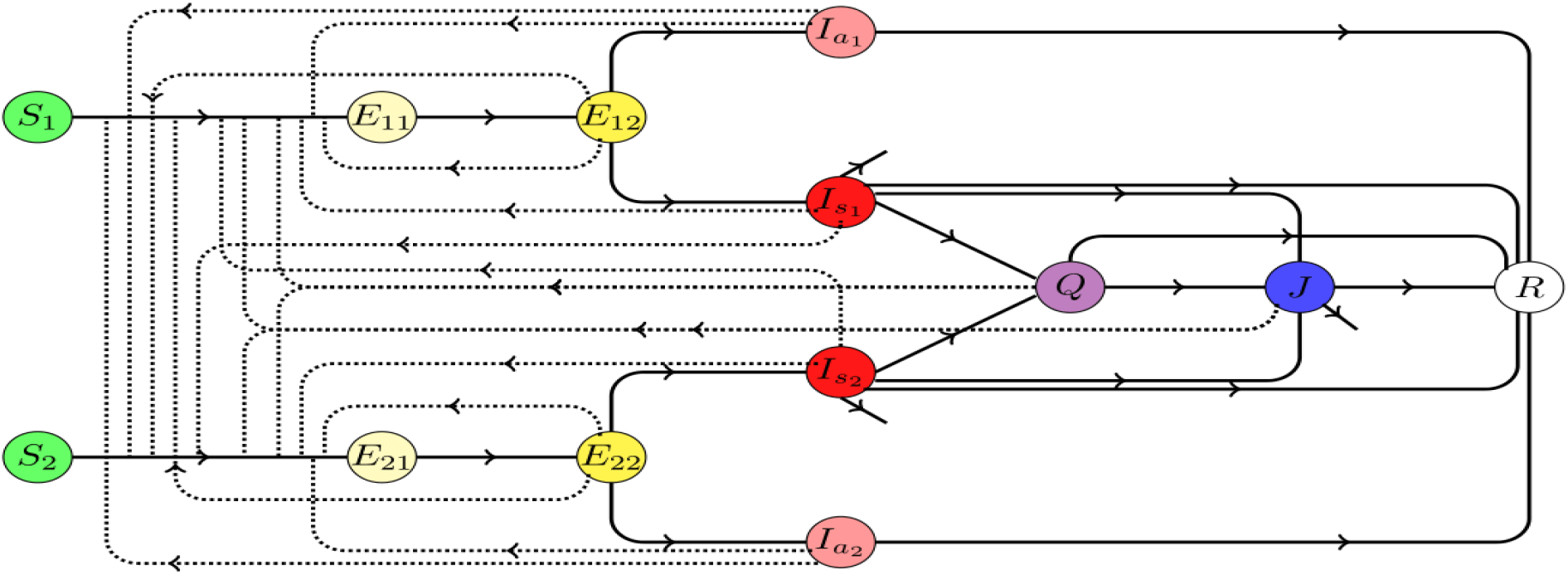
Schematic diagram for the disease progression described by the model (33). Solid arrows represent the transfer from one compartment to another while the dotted line with arrow denote the compartments responsible for disease transmission.

Before analyzing the above model, first we explain how the model (1) can be derived from the model (33) under suitable assumptions. For this purpose first adding the equations (33e) and (33f), we find

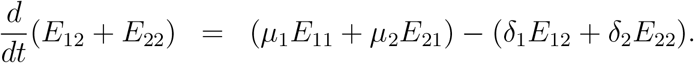

If we assume *μ*_1_ = *μ*_2_ = *μ* and *δ*_1_ = *δ*_2_ = *δ*, then we find

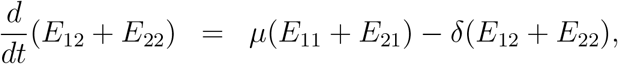

which is equation (1c) if we write (*E*_11_ + *E*_21_) = *E*_1_ and (*E*_12_ + *E*_22_) = *E*_2_ respectively.

Similarly, with the previous assumptions on parameters and variables, and additional assumptions *σ*_1_ = *σ*_2_ = *σ*, *η*_1_ = *η*_2_ = _ and *I_a_*_1_ + *I_a_*_2_ = *I_a_*, we can derive (1d) by adding (33g) and (33h). We can also derive (1e) from the equations (33i) and (33j) using the similar procedure.

Derivation of equation (1a) by adding equations (33a) and (33b) is little bit tricky. Adding (33a) and (33b), and rearranging the terms, we can write

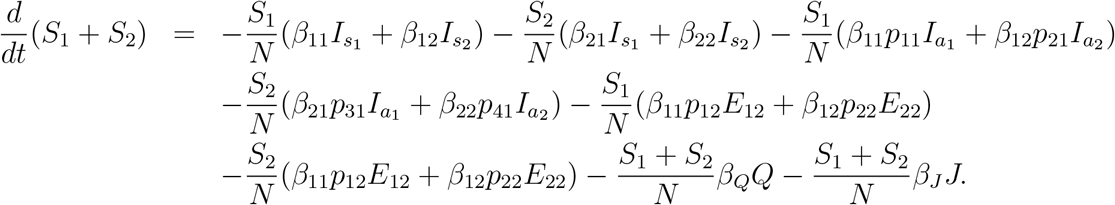

Firstly, if we assume *β*_11_ = *β*_12_ = *β*_21_ = *β*_22_ = *β*, *β*_Q_ = *p*_3_ *β*, *β*_J_ = *p*_4_*β*, then we can write

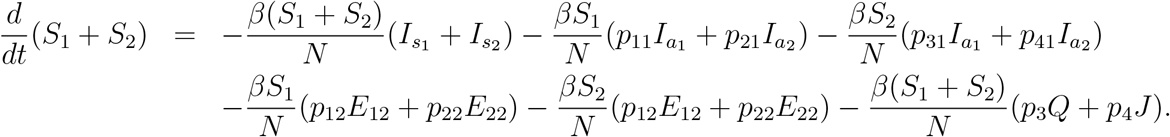

Further if we assume *p_j_*_1_ = *p*_1_, *p_j_*_2_ = *p*_2_, *j* = 1; 2; 3; 4, we can write from above equation

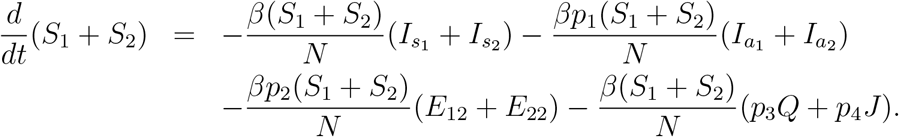

Finally writing *S*_1_ + *S*_2_ = *S*, and using previous assumptions related to *E*_2_, *I_a_* and *I_s_*, we can write above equation as

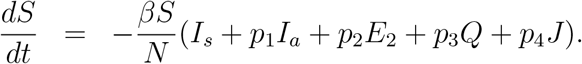

With the other necessary assumptions on the remaining parameters and variables we can derive the model (1) from (33). Once the two group of individiuals are non-distinguishable, then the model (1) results in from the model (33).

### 3.1 Controlled reproduction number

For simplicity of forthcoming calculations, we can write the first two equation (33a) - (33b) into the matrix form as follows:

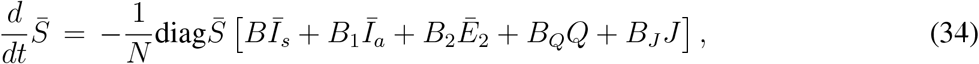

where

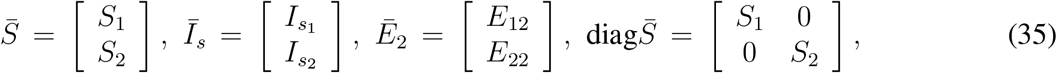

and

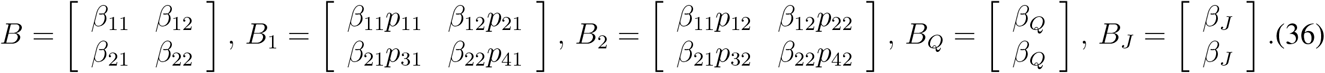

Using above approach, we can write (33c) - (33d) into a compact form as follows

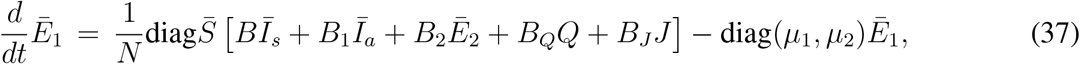

where

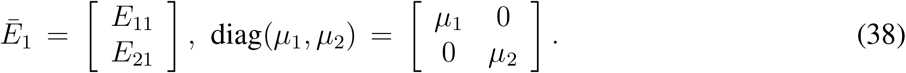

Similarly, equations (33e) - (33f), (33g) - (33h) and (33i) - (33j) can be written as

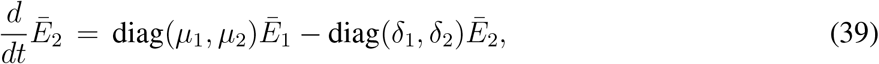

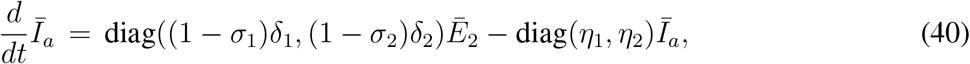

and

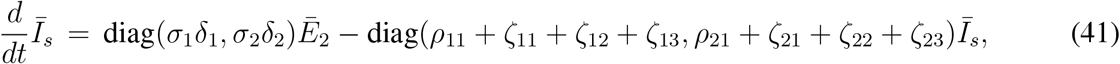

respectively.

In order to calculate the controlled reproduction number for the model (33), we follow the similar approach as we have used for the model (1). Without giving much description, here we just define the relevant matrices as follows,

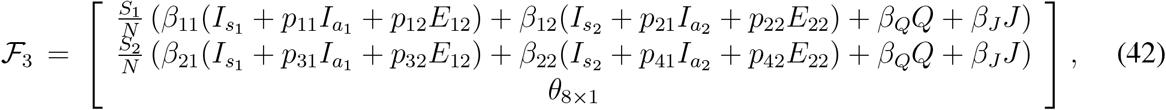

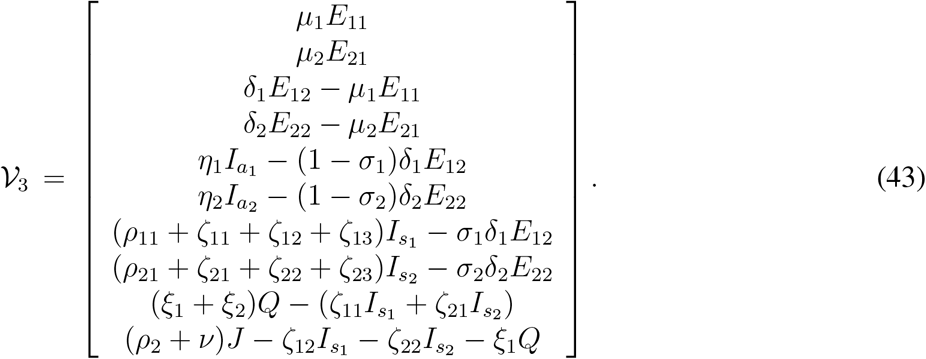

Evaluating the Jacobian matrices, at the disease free equilibrium point (*N*_1_, *N*_2_,0,0,0,0,0,0,0,0,0,0,0), corresponding to 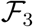 and 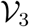, we find

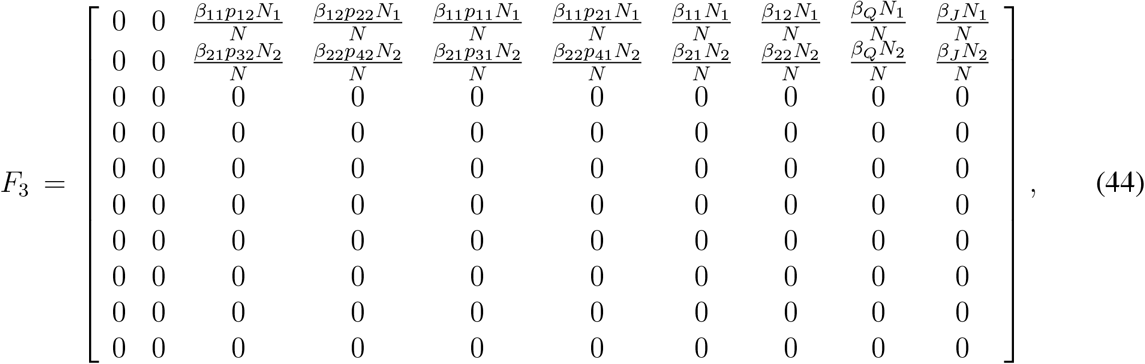

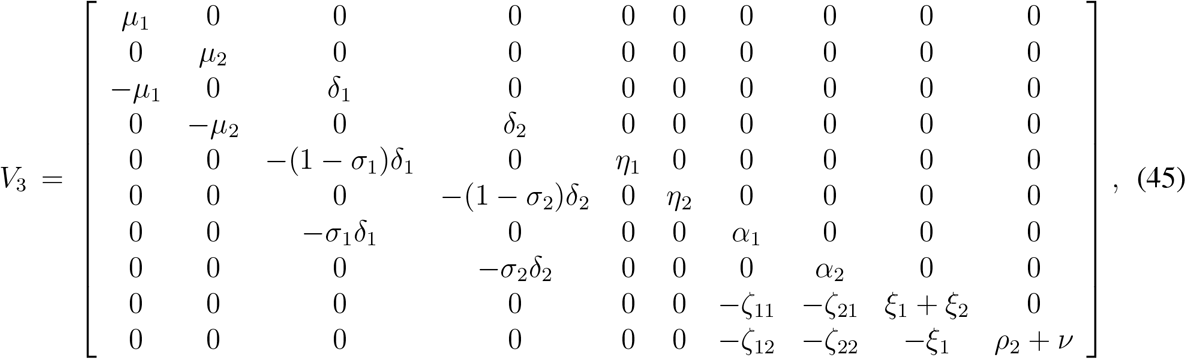

where

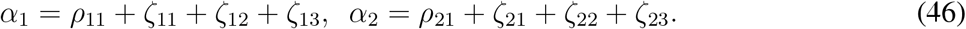

The controlled reproduction number is the largest eigenvalue of the matrix *F*_3_ *V*_3_^−1^

In terms of the matrices introduced above, we can rewrite *F*_3_ and *V*_3_ as partitioned matrices analogous to the matrices *F*_2_ and *V*_2_ as in the previous section. *F*_3_ and *V*_3_ can be rewritten as

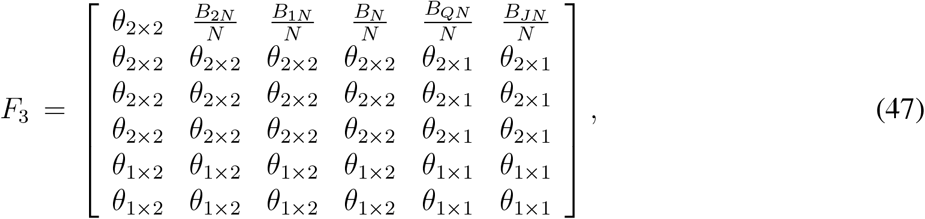

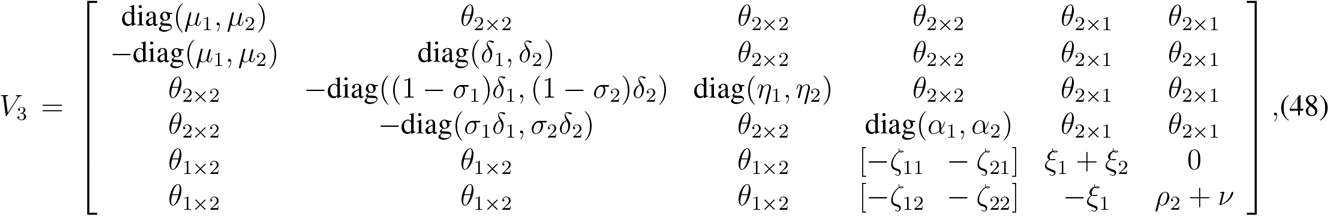

where

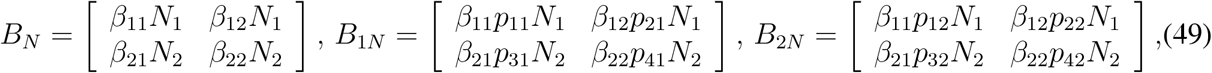

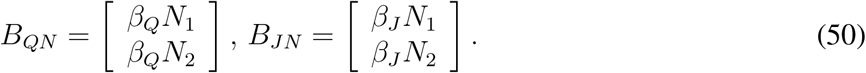

The matrix *V*_3_^−1^ is a lower triangular matrix. Based upon the non-zero entries of *F*_3_, we need the elements in first two columns of the matrix *V*_3_^−1^ in order to calculate the controlled reproduction number. If we write *V*_3_^−1^ in the following form

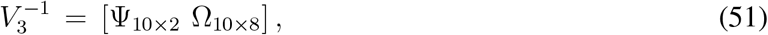

where

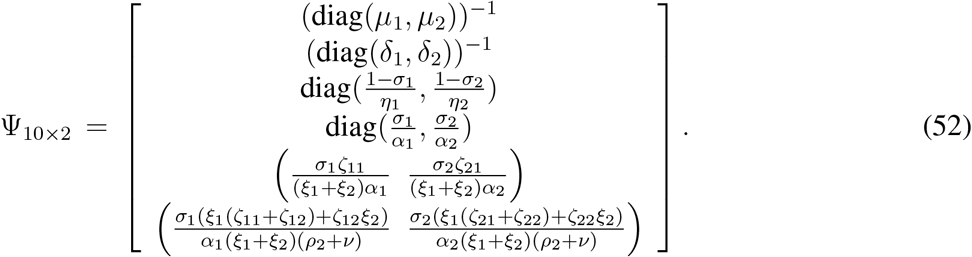

Once we calculate the matrix *F*_3_ *V*_3_^−1^, it comes out to be a matrix of following form

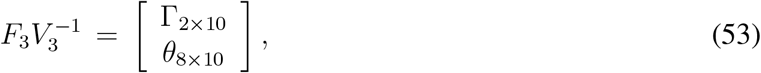

and hence the the non-zero eigenvalues can be determined from the first 2 × 2 block of the block matrix Γ_2×10_ involved with *F*_3_ *V*_3_^−1^. The entries of the first 2 × 2 block can be calculated as follows,

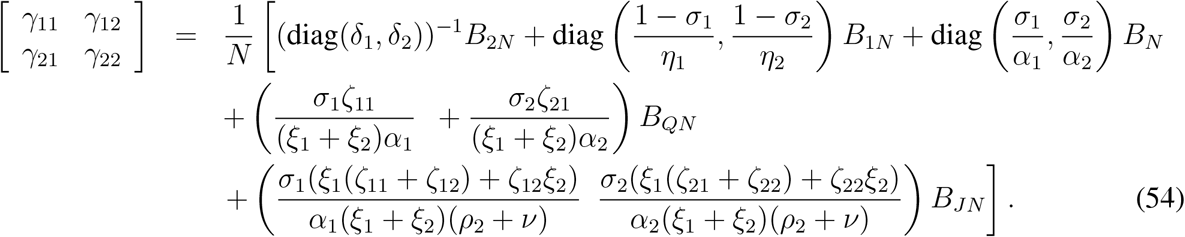

Explicit expressions for *γ_ij_*, (*i*, *j* = 1, 2) are given by

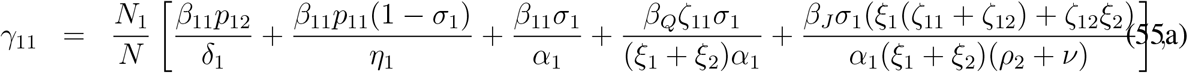

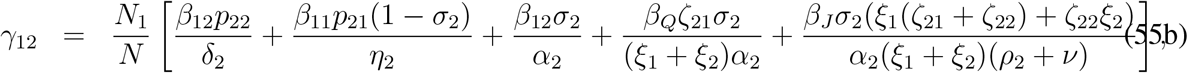

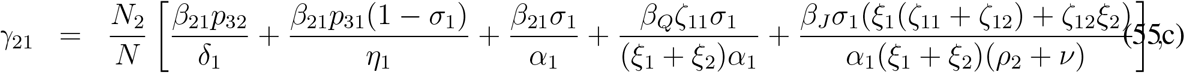

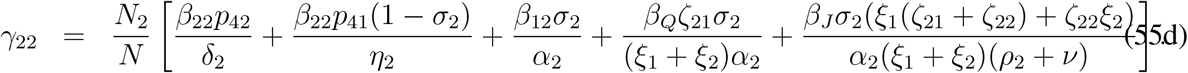

The matrix *F*_3_ *V*_3_^−1^ has at most two non-zero eigenvalues and eight zero eigenvalues. The maximum positive eigenvalue of the matrix *F*_3_ *V*_3_^−1^ is the controlled reproduction number for the model (33). As a matter of fact the controlled reproduction number for the model (33) is the largest eigen-value of the matrix [*γ_ij_*]_2×2_ and is given by

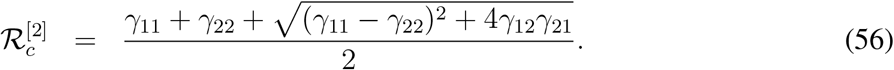

Here the superscript ‘[2]’ stands for the second mathematical model considered in this manuscript, that is for the model (33).

### 3.2 Maximum size of epidemic

Derivation of maximum size of the epidemic is tedious but can be obtained through step by step calculations. From (33a) & (33b), integrating between the limits *t* = 0 and *t* = *t_f_*, we find

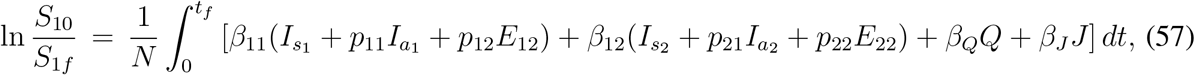

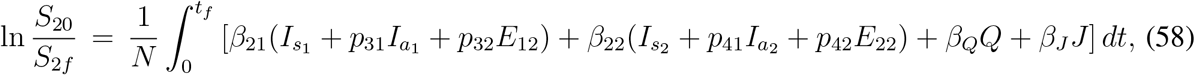

where *S_j_*_0_ and *S_jf_* denote initial and final size of the *j*-th susceptible class, *j* = 1,2. Now we assume that the model (33) is subjected to the initial conditions such that *S*_10_, *S*_20_, *E2*_110_, *E*_210_ > 0 and all other components are absent at the initial time point *t* = 0. Consequently *S*_10_ + *S*_20_ + *E*_110_ + *E*_210_ = *N*. Now integrating (33e), (33f) between *t* = 0 and *t* = *t_f_* and with the assumption that *E*_120_ = *E*_220_ = *E*_12_*_f_* = *E*_22_*_f_* = 0, we find

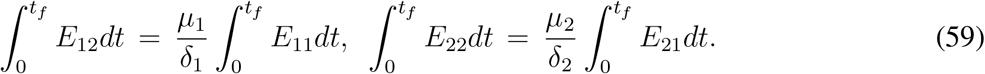

Next integrating (33g), (33h) between *t* = 0 and *t* = *t_f_* and with the assumption that 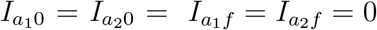, using (59) we find

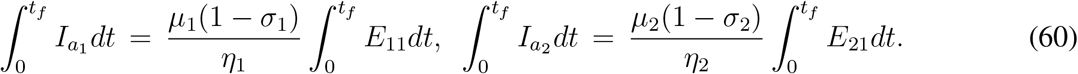

Proceeding in a similar way and using the results in (60), we get from (33i), (33j)

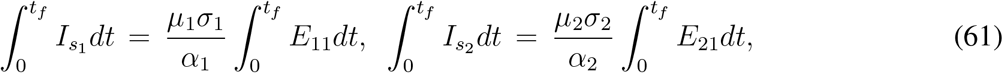

where *α*_1_ and *α*_2_ are defined in (46). Using the above result, from (33k) we get

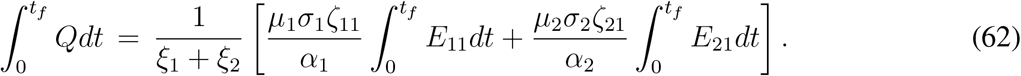

Finally, integrating (33l) between *t* = 0 and *t* = *t_f_* and using *J*_0_ = 0 = *J_f_*, we can write

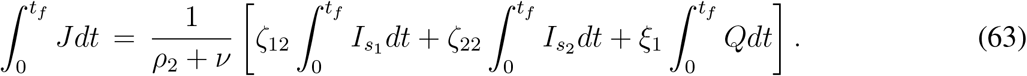

Using the results from (62) and (63), we can express the 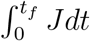 *Jdt* in terms of 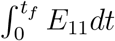 *E*_21_*dt* and 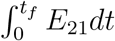 *E*_21_*dt*. Now adding the equations (33a) and (33c) and then integrating between *t* = 0 and *t* = *t_f_*, we find

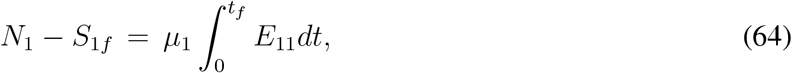

where we have used *E*_110_ = *E*_11_*_f_* = 0 and *S*_10_ = *N*_1_. Similarly, adding the equations (33b) and (33d) and following the same approach, we can find

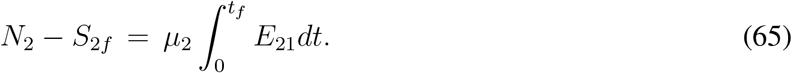

Now using the results (59) – (63), we get from (57),

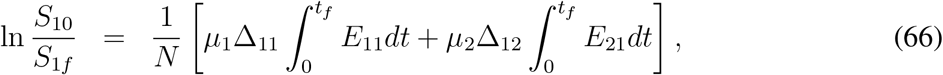

where

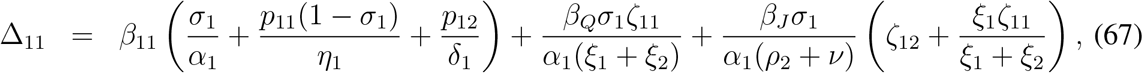

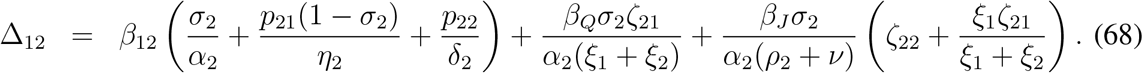

Proceeding in a similar way, from (58), we can derive

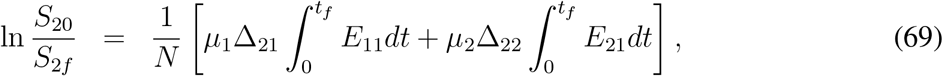

where

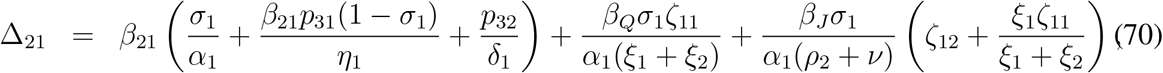

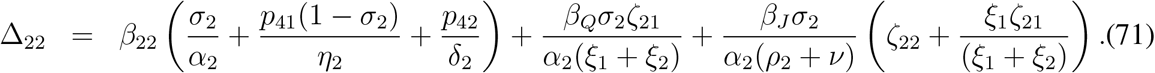

Eliminating 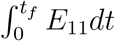 *E*_11_*dt* and 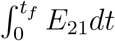 *E*_21_*dt* between (64), (65) and (69), we find the following two equations

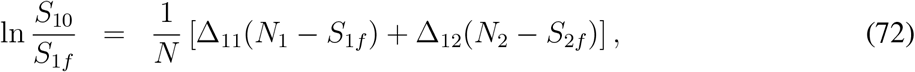

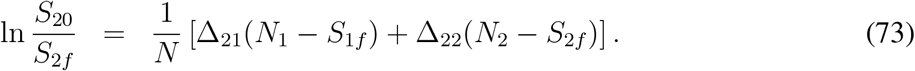

Without any loss of generality, writing *S*_10_ = *N*_1_ and *S*_20_ = *N*_2_ and introducing the notations 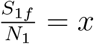, 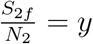 we get from above system equations

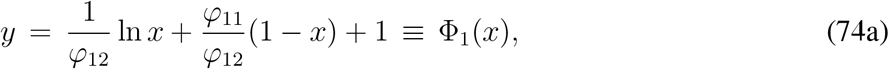

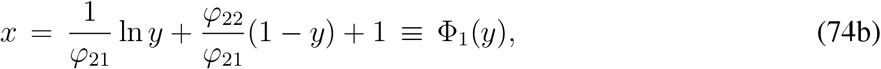

where

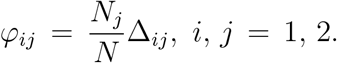

It is interesting to note that the following two matrices are similar matrices,

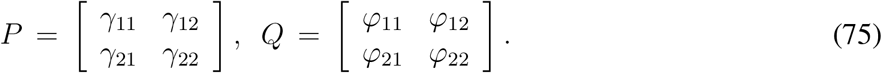

If we define a diagonal matrix

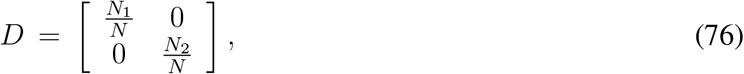

then we can verify that

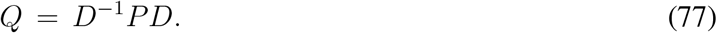

Entries of both the matrices *P* and *Q* are positive and as they are similar, the spectrum of two matrices are same. Hence the largest eigenvalue of the matrix *Q* is equal to the largest eigenvalue of the matrix *P* that is equal to 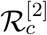.

The existence of final size of the epidemic depends upon the existence of a point of intersection between two curves *y* = *ϕ*_1_(*x*) and *x* = Φ_2_(*y*) within the unit square [0,1] × [0,1]. Existence of such point of intersection follows from the Th. 1 in [37]. The two curves *y* = *ϕ*_1_(*x*) and *x* = Φ_2_(*y*) intersect each other at a point 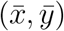, 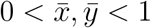 whenever 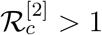.

## 4 Numerical simulations

In this section we present the numerical simulation results for two models (1) and (33) considered in this work. First we present the simulation results for the model (1) by fitting the numerical simulation results with the COVID-19 data for Germany as an example. All the data used in this manuscript are taken from [3]. In order to ensure that the obtained result and the validity of the model is not country specific rather it can be matched with the data for other countries. Fitting of the data for Italy, UK and Spain are provided at the appendix.

The sensitivity of various parameters involved with the model (1) plays an important role behind numerical simulations. It would be quite lengthy calculations if we want to perform the same for the model (33) and hence we have restricted ourselves to the first model only. For numerical simulations and fitting with the data, we have considered the rate of infection as a function of time and hence some of the estimated parameters do not remain fixed throughout the simulation and hence we have presented the sensivity indices with respect to the estimated parameter values.

### 4.1 Sensitivity analysis

The sensitivity analysis for the endemic threshold (the controlled reproduction number 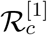) in equation (15) tells us how important each parameter is to disease transmission. It is used to understand parameters that have a high impact on the threshold 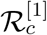 and should be targeted by intervention strategies. More precisely, sensitivity indices allows us to measure the relative change in a variable when a parameter changes. For that purpose, we use the normalized forward sensitivity index of a variable with respect to a given parameter, which is defined as the ratio of the relative change in the variable to the relative change in the parameter. If such variable is differentiable with respect to the parameter, then the sensitivity index is defined as follows.

The normalized forward sensitivity index of 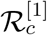, which is differentiable with respect to a given parameter *θ* (say), is defined by

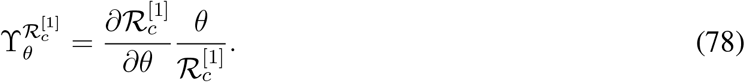

From direct calculation and without using the numerical values we can determine the signs of sensitivity indices with respect to some of the parameters. For the remaining sensitivity indices, we need to take help of the numerical examples. The normalized forward sensitivity indices of 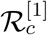 with respect to the parameters which are found to be clearly positive are given by,

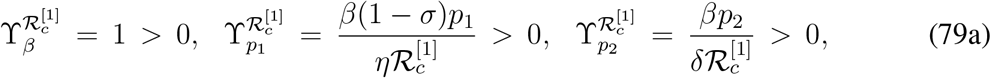

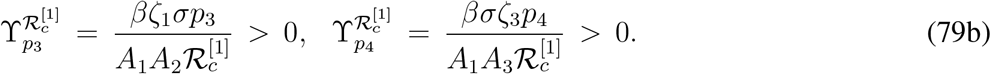

The normalized forward sensitivity indices of 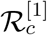 with respect to the parameters which are found to be clearly negative are given by,

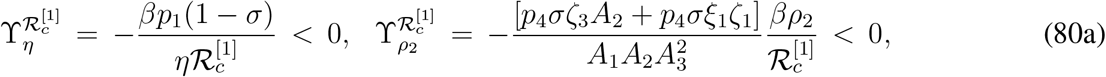

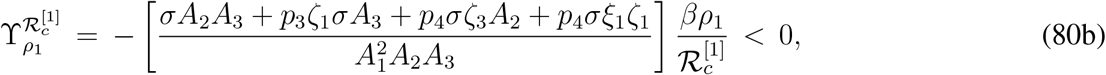

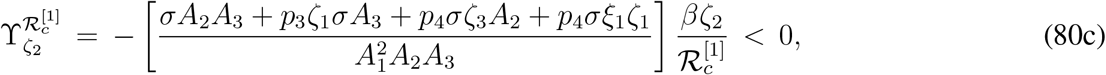

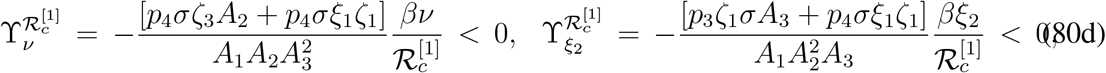

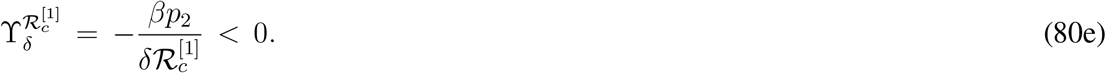

The signs of normalized forward sensitivity indices of 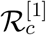 with respect to rest of the parameters can not be determined from their explicit expressions and we need to take help of some numerical examples. Such normalized forward sensitivity indices are as follows,

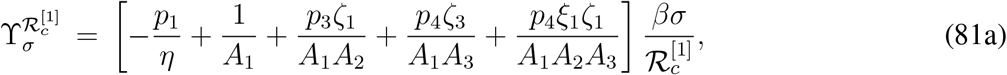

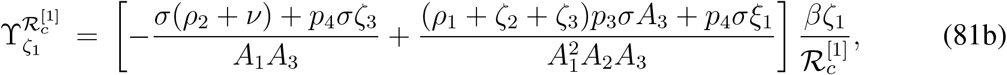

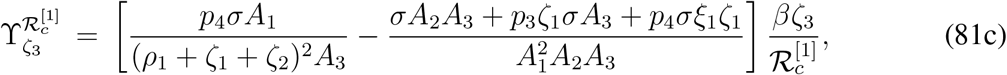

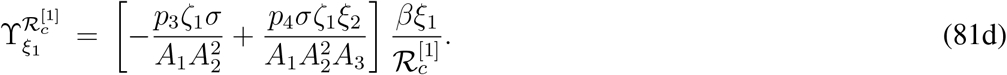

We use the sensitivity indices to understand parameters that have a high impact on 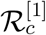. The values of the sensitivity indices for different parameters depend on the choice of parameter values. As we have mentioned earlier, the values of the sensitivity indices can be positive or negative. The most positive (or negative) value of the sensitivity index for a parameter indicates that parameter is most sensitive to 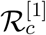 and the least positive (or negative) value of the sensitivity index for a parameter indicates that parameter is least sensitive to 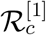.

**Table 2:** Initial parameter values used as the initial guess for the (1) and sensitivity indices of 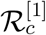 with respect to the parameters and values mentioned here.

| Parameter | Values | Sensitivity Index |
| --- | --- | --- |
| $\beta$ | varying | 1 |
| $p_1$ | 0.2 | 0.3340 |
| $p_2$ | 0.2 | 0.2969 |
| $p_3$ | 0.2 | 0.0158 |
| $p_4$ | 0.2 | 0.0634 |
| $\delta$ | 0.75 | -0.2969 |
| $\sigma$ | 0.1 | 0.3321 |
| $\eta$ | 0.6 | -0.3340 |
| $\rho_1$ | 0.1 | -0.09 |
| $\rho_2$ | 0.07 | -0.0477 |
| $\zeta_1$ | 0.07 | 0.0348 |
| $\zeta_2$ | 0.1 | -0.09 |
| $\zeta_3$ | 0.14 | 0.0417 |
| $\xi_1$ | 0.14 | -0.0015 |
| $\xi_2$ | 0.1 | -0.0143 |
| $\nu$ | 0.05 | -0.0341 |

The sensitivity index may depend on several parameters of the system, but also can be constant, independent of any parameter. For example, 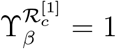 means that increasing (decreasing)*β* by a given percentage increases (decreases) always 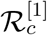 by that same percentage. The estimation of a sensitive parameter should be carefully done, since a small perturbation in such parameter leads to relevant quantitative changes. On the other hand, the estimation of a parameter with a rather small value for the sensitivity index does not require as much attention to estimate, because a small perturbation in that parameter leads to small changes. From Table 2, we conclude that the most sensitive parameters to the basic reproduction number 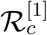 of the COVID-19 model (1) are *β*; *p*_1_; *p*_2_; *η*; *σ*; *δ*. In concrete, an increase of the value of *β*; *p*_1_; *p*_2_; *σ* will increase the basic reproduction number by 100%, 33:4%, 29:69%, 33:21% respectively and an increase of the value of *η*; *δ* will decrease 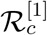 by 33:4% and 29:69% respectively. Sensitivity indices with respect to the parameter values as presented in Table 2 can be visualized from Fig. 4.

**Figure 4:**
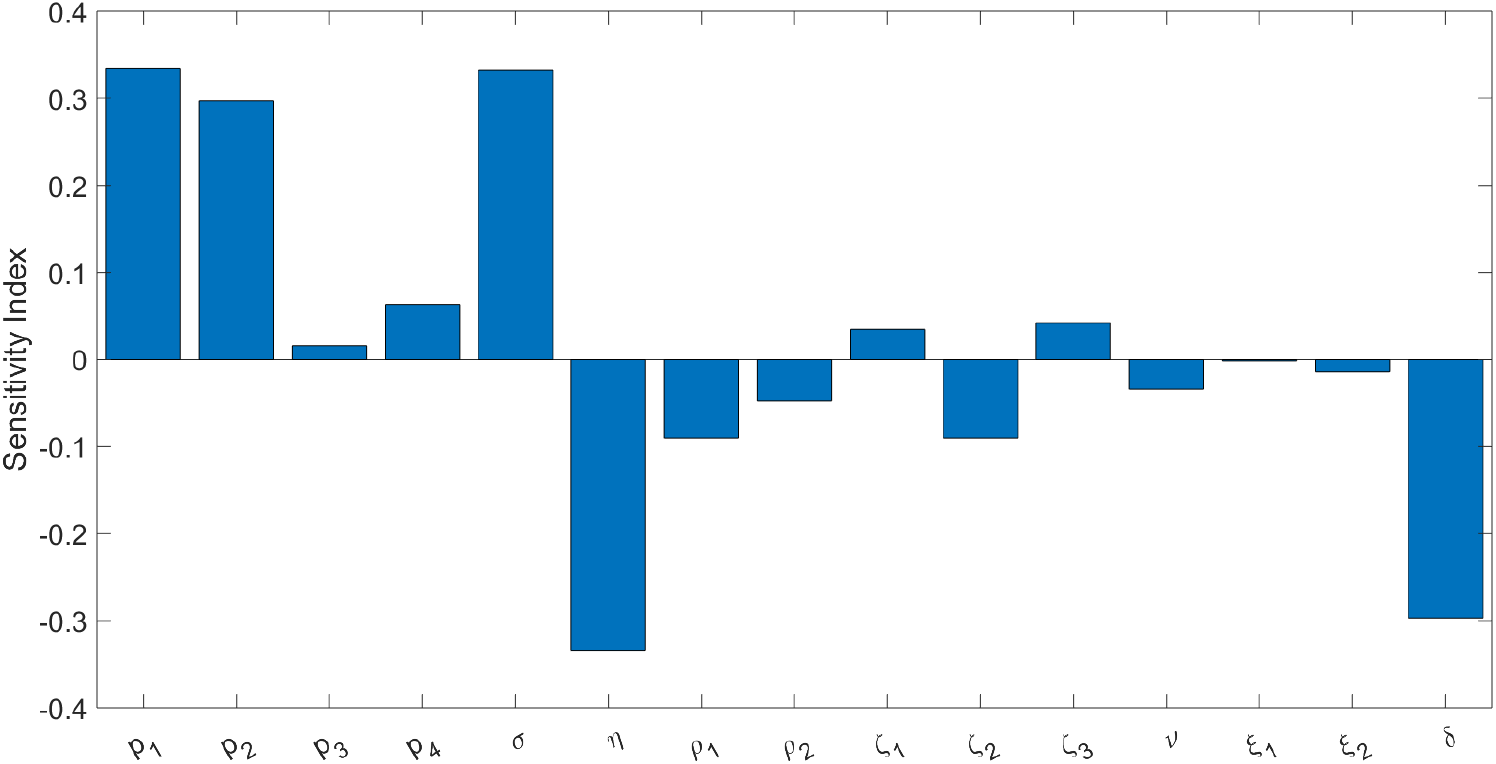
Sensitivity indices of 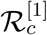 with respect to the parameters involved with the model (1) and their reference values as mentioned in Table 2.

### 4.2 Simulation results for model - 1

We have fitted the parameters *β*; *p*_1_; *p*_2_; *p*_4_; *μ*; *δ*; *σ*; *η*; *ρ*_1_; *ξ*_2_ for the range of the values given in Table 1 and took *ζ*_1_; *ζ*_2_; *ζ*_2_; *ξ*_2_; *ρ*_2_; *ν* fixed as given in Table 1. We estimated those values for three different time intervals (1-30 th day, 31-42 nd day, 43-95 th day). We have considered the initial values of the parameters as given in Table 2. We have chosen the time interval and end values of different compartments from the fittings over the previous time intervals.

The optimization of parameters to describe the outbreak of COVID-19 in Germany was fitted by minimizing the Sum of Squared Errors (SSE), in such a way that the solutions obtained by the model approximate the reported cumulative number of infected cases. We applied three searches to minimize the SSE function: by using a gradient-based method first, followed by a step of minimization with a gradient-free method, again followed by a third step of gradient-based method. MATLAB based nonlinear least-square solver *fmincon* and *patternsearch* are used to fit simulated and observed daywise cumulative number of infected cases for three different time intervals. Detailed description of this method and its implementation can be found in [32, 38–40].

The model (1) is simulated with best fitted parameter values as mentioned above and the simulation result against the daily reported data and cumulative data are presented in Fig. 5a. In this case the values of *β* are constants over three different range of days starting from the initial date of COVID-19 epidemic spread in Germany. For this figure some of the parameters are estimated as described above. Another simulation result is presented in Fig. 5b where initially the value of *β* is constant for several days, and then monotone decreasing over a range of days and then remain constant for the rest of the period. The variation of *β* with respect to time is shown in Fig. 6. Interestingly the data are fitted well in the case of time varying *β* where *β*(*t*) is continuous. For the simulation result presented in Fig. 5b, the number of days for which *β*(*t*) remains constant and then start decaying are adjusted in such a way that the outcome matches well with both the cumulative and daily data. Accordingly the slope of decaying *β*(*t*), the day up to which it decays and the lower value of *β*(*t*) are chosen. The numerical simulation and fitting with the data is carried out with the objective that the simulation result should be pretty close to the cumulative number of reported cases. We can claim that our attempt is successful as the cumulative number of reported cases for Germany on 15th May, 2020 obtained from the simulation is 180,788 and the reported data (see [3]) shows it is equal to 182,250. In this case the choice of *β*(*t*) is as shown in Fig. 6a. Cumulative number of reported cases obtained from the simulation with the *β*(*t*) as shown in Fig. 6b is equal to 180,808. Both the simulated values are close to the reported data. Relative percentage error is less that 1% and is precisely equal to 0:79%.

**Figure 5:**
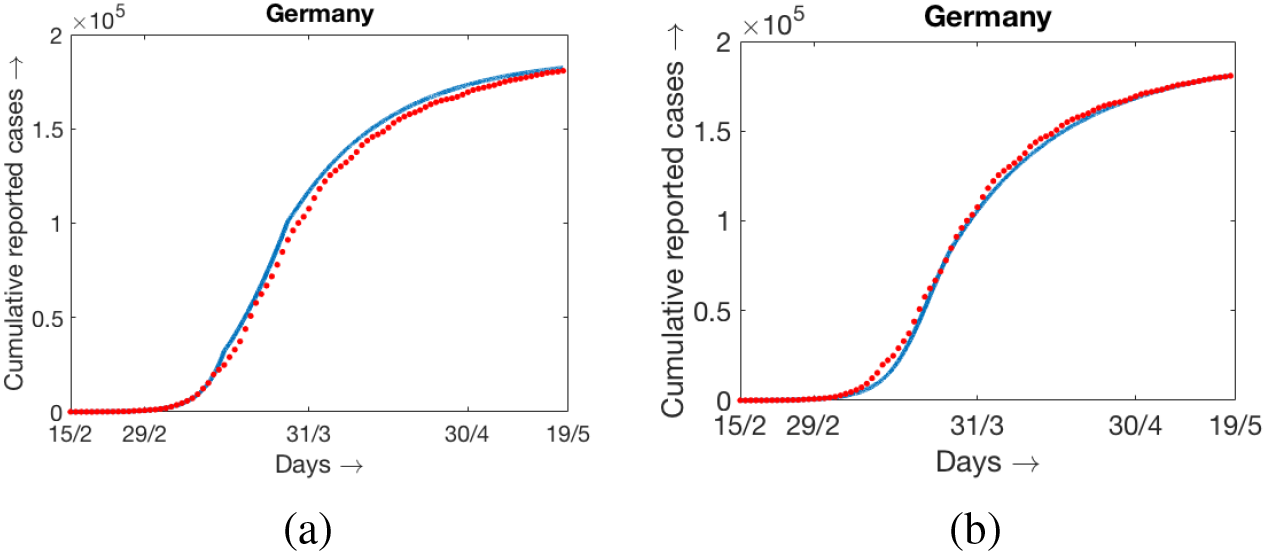
Blue curves indicate the model simulation and the red dotted curves indicate the reported data for cuculative infected population. Simulation results are obatined for two different forms of *β*(*t*), (a) with *β*(*t*) as shown in Fig. 6a; (b) with *β*(*t*) as shown in Fig. 6b.

**Figure 6:**
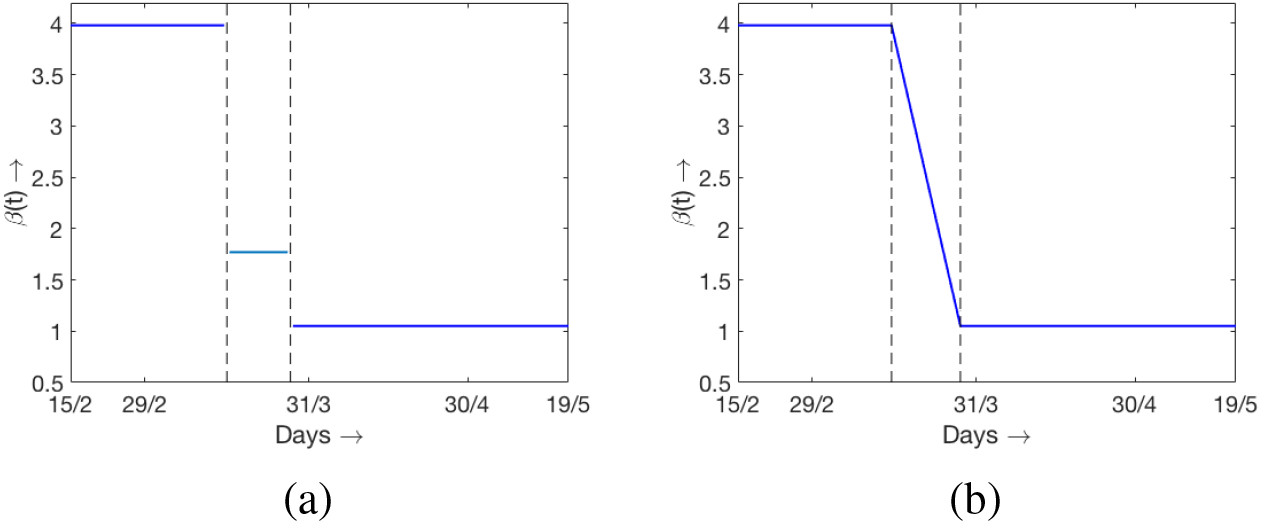
The values of *β* changing with time *t* and used in (a) Fig. 5a and (b) Fig. 5b respectively.

Simulations and fitting are done with the reference to cumulative number of reported cases however the obtained results are also in good agreement with the daily data also. As there is no uniformity of the daily reported cases, may be due to irregular reporting, we have used 7 days moving agerage as daily data instead of daily raw data. Simulation results with two different types of *β*(*t*) and 7 days moving average for daily reported data are shown in Fig. 7. The estimated and other parameter values for the simulation with *β*(*t*) is constant over three different time intervals are presented in Table 3 along with the sensitivity incides.

**Figure 7:**
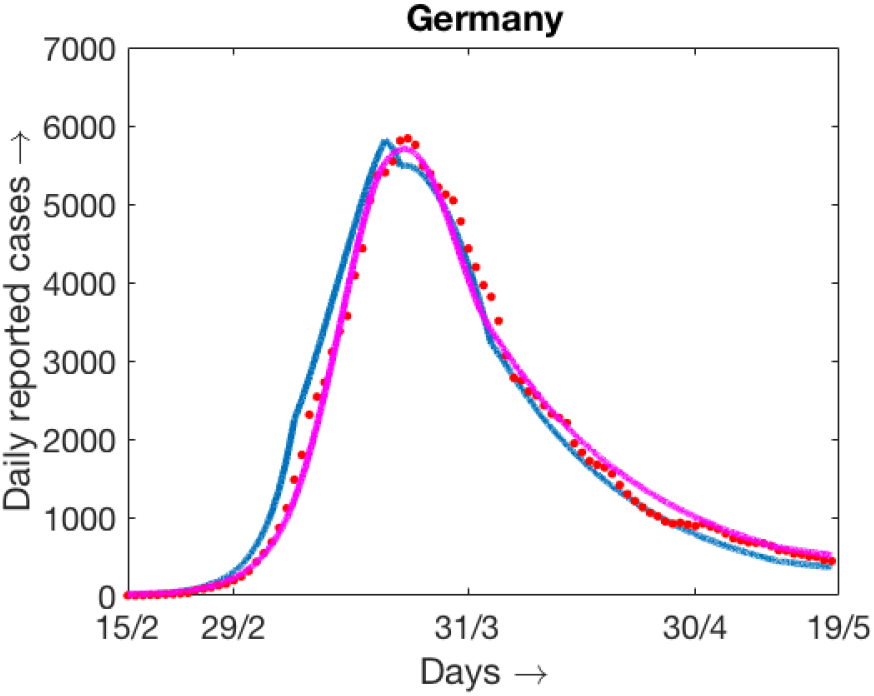
Simulation results and daily reported cases (7 days moving average) for two different form of *β*(*t*). Blue curve and magenta curves are for *β*(*t*) as shown in Fig. 6a and Fig. 6b respectively.

**Table 3:** Best fitted values for the parameters of (1) for Germany.

| Parameter | Best fit values<br>for 1st 30 days<br>(Sensitivity Index) | Best fit values<br>for next 12 days<br>(Sensitivity Index) | Best fit values<br>upto 95th day<br>(Sensitivity Index) |
| --- | --- | --- | --- |
| $\beta$ | 3.98 (1) | 1.77 (1) | 1.05 (1) |
| $p_1$ | 0.177 (0.2383) | 0.16 (0.2204) | 0.34 (0.5099) |
| $p_2$ | 0.3 (0.4039) | 0.3 (0.4133) | 0.05 (0.0914) |
| $p_3$ | 0.05 (0.0048) | 0.05 (0.0049) | 0.05 (0.0053) |
| $p_4$ | 0.05 (0.0192) | 0.05 (0.0196) | 0.05 (0.0213) |
| $\delta$ | 1 (-0.4039) | 1 (-0.4133) | 0.82 (-0.0914) |
| $\sigma$ | 0.1 (0.3314) | 0.1 (0.3418) | 0.1 (0.342) |
| $\eta$ | 0.9 (-0.2383) | 0.9 (-0.2204) | 0.9 (-0.5099) |
| $\rho_1$ | 0.1 (-0.0873) | 0.1 (-0.0893) | 0.1 (-0.0972) |
| $\rho_2$ | 0.07 (-0.0144) | 0.07 (-0.0148) | 0.07 (-0.0161) |
| $\zeta_1$ | 0.07 (-0.0067) | 0.07 (-0.0069) | 0.07 (-0.0075) |
| $\zeta_2$ | 0.1 (-0.0873) | 0.1 (-0.0893) | 0.1 (-0.0972) |
| $\zeta_3$ | 0.14 (-0.0715) | 0.14 (-0.0732) | 0.14 (-0.0796) |
| $\xi_1$ | 0.14 (-0.00047) | 0.14 (-0.00048) | 0.14 (-0.00052) |
| $\xi_2$ | 0.1 (-0.0043) | 0.1 (-0.0044) | 0.1 (-0.0048) |
| $\nu$ | 0.05 (-0.0103) | 0.05 (-0.0105) | 0.05 (-0.0115) |
| $\mu$ | 0.28 (—) | 0.28 (—) | 0.28 (—) |

Consideration of changing *β* over different range of durations is an important observation of our present work. It is important to mention that the number of intervals over which we have to take different values of *β* is not always limited to three as in the case of Germany. The number of intervals over which we need to choose different values of *β* in order to match the simulation results with the data from different countries varies significantly. Of course, in all such simuations the values of *β* are decreasing with respect to the number of days. The simulations results for three more countries, namely Italy, UK and Spain, are provided in the appendix along with the result for Germany again where the 95% confidence intervals are shown. The parameter values used, the range of days over which *β* remains constant and estimated parameter values are also provided at the appendix.

### 4.3 Simulation results for model - 2

In order to understand the usefullness of the model (33), we now present some numerical simulation results. In Fig. 5 we have presented the simulation results for the model (1) up to 19th of May. For the fitting with data, we have obtained three different values of *β* as *β*_1_ = 3.98, *β*_2_ = 1.77 and *β*_3_ = 1.05. The partial lockdown was started at Germany on 13th March, 2020 and then went into complete lock lockdown step by step. The process of unlocking at the essential sectors were started from 15th of May, 2020 and by that time the spreading speed of COVID-19 was much reduced. There are several restrictions like using mask, maitaining social distances and many other restrictions are in place in order to prevent the further spread of the disease. As of now the disease is not completely eradicated rather a minor number of cases are getting reported on a regular basis but the situation is under control.

We are interested to see the situation if all the citizens do not follow the suggested restrictions then what can happen afterward. For this purpose we now perform an interesting numerical simulations. We have simulation results for the model (1) as described at the previous subsection up to 19th of May. Now we start simulating the model (33) from the day immediately after 19th May and continue up to the end of November 2020. We assume that a fraction of population are not following the guideline and other fraction is following the safety and preventive norms appropriately. Let us define the “coefficient of social interactions” as *S*_1_/*N* and is denoted by K. If we assume that the 10% of the population is not following the norm then *S*_1_/*N* ≡ *K* = 0.1 and hence *S*_2_/*N* = 0.9 = 1 − *K*. We assume that *β*_11_ = *β*_1_, *β*_22_ = *β*_3_ and *β*_12_ = *β*_21_ = (*β*_1_ + *β*_3_)/2 where *β_j_*, (*j* = 1, 2, 3) are the values determined by fitting the data with the first model. Initial densities of *E*_11_ and *E*_21_ can be distributed with the same proportion to *K* and 1 − *K* of the value of *E*_1_ on 19th May. Same procedure is adopted for other components except *Q*, *J* and *R*. For simplicity we also assume that the other parameters involved with the two group model are equal to the values of the corresponding parameter involved with the model (1) and is equal to the numerical values as mentioned at the last column of Table 3. For clearer understanding we can mention here some of the parameter values: *P_j_*_1_ = *p*_1_ = 0.34 (*j* = 1, 2, 3, 4), *σ*_1_ = *σ*_2_ = *σ* = 0.1 and so on.

With above choice of initial values and parameter values now we simulate the model (33) up to the end of November, 2020. Two different simulations are performed and the simulation results are presented in Fig. 8. For first simulation we have considered *K* = 0.1 fixed up to the end of November 2020 (see Fig. 8a). Second simulation is performed for *K* = 0.1 upto the end of August and then *K* = 0.15 during September to November 2020. In the figure we have ploted the consolidated compartments *E*_1_, *E*_2_, *I_a_* and in order to avoid any confusion where we have obtained the values of these compartments by adding the values of the respective compartments in two group. To be specific, *E*_1_ = *E*_11_ + *E*_12_ and similary for other compartments. A relatively small increase of *K* lead to an essential acceleration in the disease progression.

With the parameter setup mentioned above one can calculate the controlled reproduction number 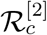 using the detailed formula given in Sub-section 3.1. Without any loss of generality we can make a crude assumption that 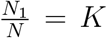 in order to find a value of 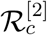. Clearly the measure for 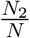 will be 1 − *K*. With *K* = 0.1, we can see slow growth in exposed and infected compartments as shown in Fig. 8a as we find 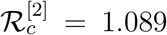. Now if we change the value of *K* to *K* = 0.15 the revised measure of 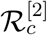 becomes 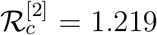. These results ensure the usefulness of our model along with the mathematical details and supportive numerical simulations.

**Figure 8:**
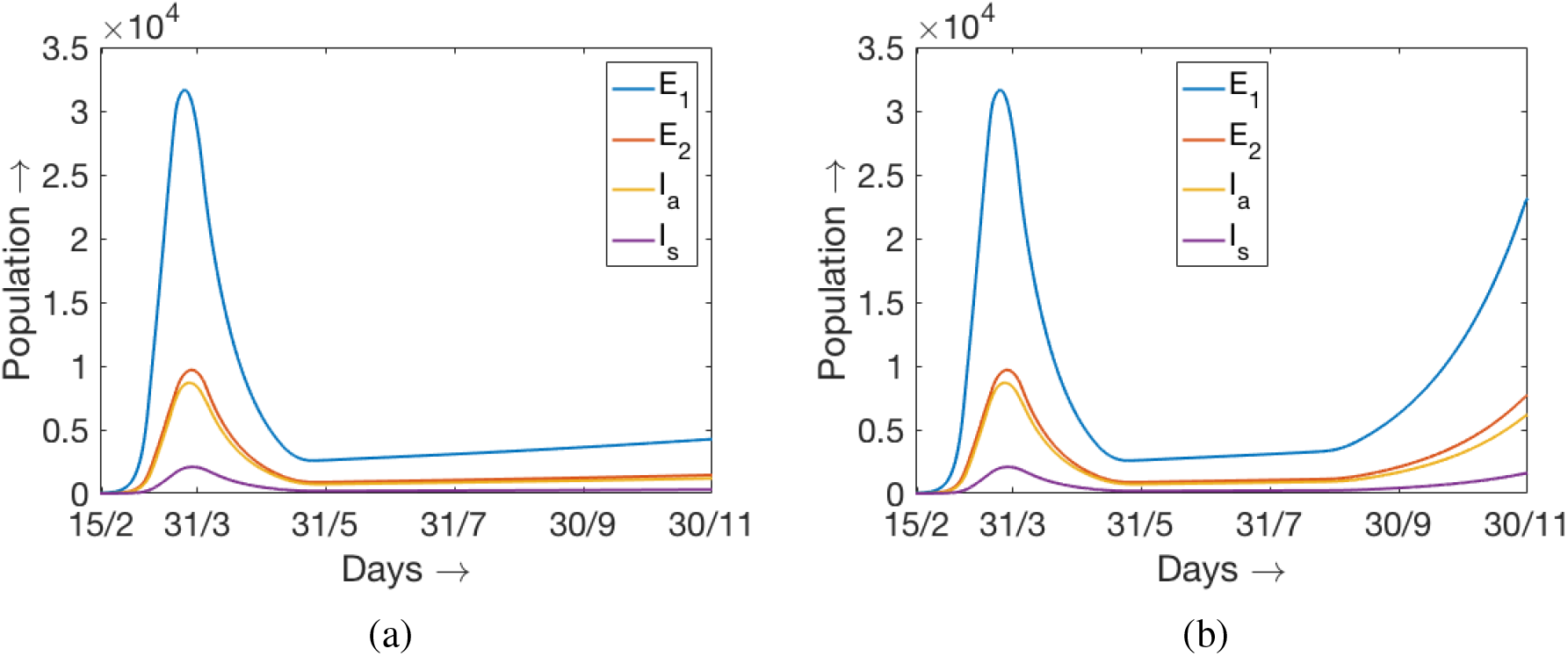
Time evolution of *E*_1_, *E*_2_, *I_a_* and *I_s_* obtained from the numerical simulation of the two models (1) and (33) from 15th February to 30th November 2020 as described in the text. Two different values of K are used for the duration 1st September to 30th November, (a) *K* = 0.1 and (b) *K* = 0.15.

## 5 Model with relapse

Most of the important features and several issues related to COVID-19 remain unclear and immense scientific efforts are required to understand various important issues. As it is reported at many sources that some of the recovered individuals can become infected after a certain period of time as the recovery is seems to be temporary. In this section we want to revisit the model (1) with the modification that a fraction of recovered population can return back to the susceptible class. We are mainly interested to calculate the final size of the infected compartment due to fact that the recovered individuals join the susceptible class after a certain number of days. In order to find a rough estimate of the size of the infected compartment and to obtain their estimate explicitly we can make a crude assumption. The assumption is that the disease related death rate is negligible. With these assumptions the model (1) with relapse of the disease can be written as follows

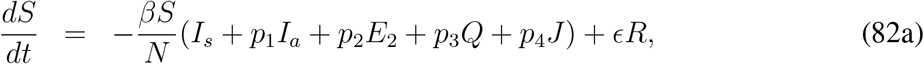

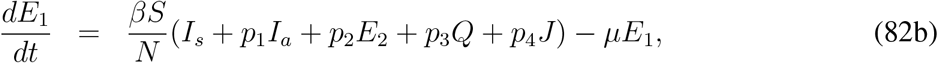

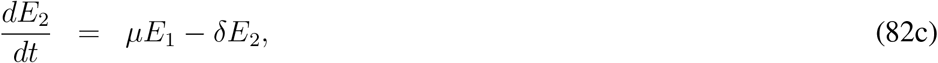

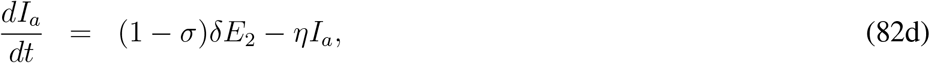

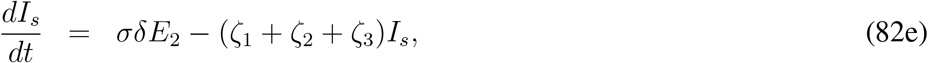

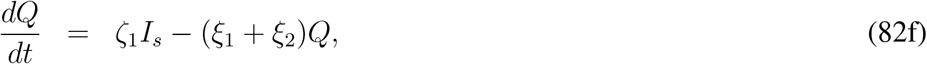

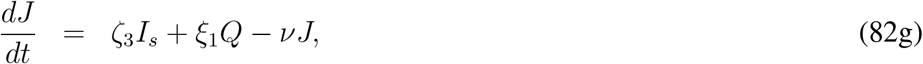

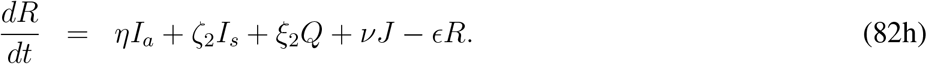

This model admits an endemic equilibrium point apart from the disease free equilibrium point. Let us denote the components of endemic equilibrium point as 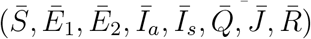, then equating the right hand sides of (82c) to (82g), we find

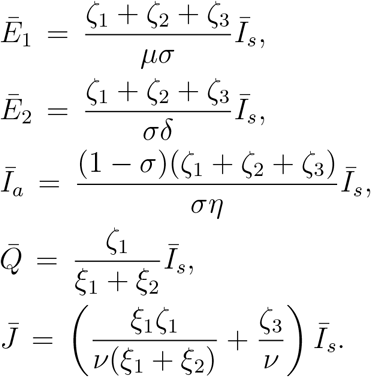

Now equating the right hand sides of (82a) and (82b) to zero and then adding, we can find

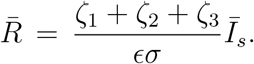

Now we consider the following equation obtained from (82b),

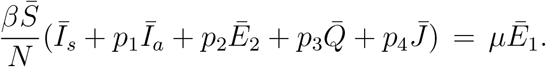

Using the expressions for 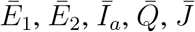 in terms of 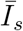, we can write

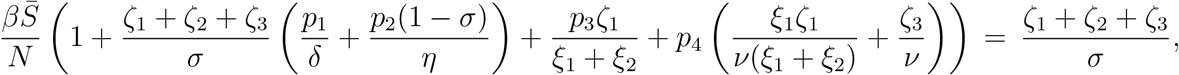

and then using 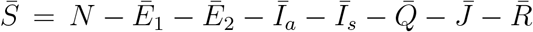 and the relevant expressions we can find

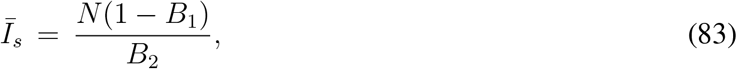

where

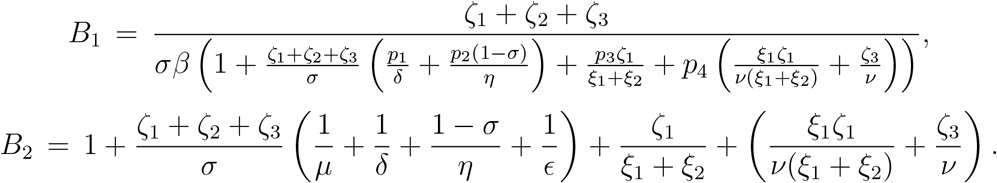

In order to come up with a closed form of estimate for the endemic steady-state in case of relapse of the disease, we have assumed that the number of COVID-19 related death is negligible. In reality this is not true however it help us to have a rough estimate for the endemic steady-state dependning upon the value of *∊*. The parameter *∊* is related to the relapse rate as 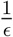 is the average number of days after which a recovered individual can have fresh infection. It is diffcult to derive explicit condition for which *B*_1_ is less than one in order to have a feasible values for the components of endemic equilibrium point in case of replase. In order to have a feasible values of *Ī_s_* and other components we should have *B*_1_ > 1. Interestingly, with the parameter values mentioned at the last column of Table 3, we can calculate *B*_1_ = 1.12 where *β* = 1.07 and note that 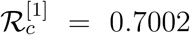. In order to have an admissible value of *B*_1_, if we consider *β* = 1.2 and with other parameter values are same as in the third column of Table 3 we find *B*_1_ = 0.98 although 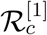 remains less than one. Finally assuming *∊* = 0.01, that is recovered individuals can join the susceptible class after 100 days on an average, we find 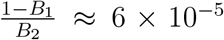. So a rough idea about the estimated number of infected individuals at the endemic state will be 6 × 10^−5^ × *N* where *N* is the total population of the country. The measure 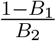 decreases with the decrease in e as we can calculate 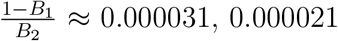 for *∊* = 0.005, 0.0033 respectively.

## 6 Discussion

A wide range of mathematical approaches are adopted to develop viable mathematical model to understand the propagation of disease spread for COVID-19. The proposed mathematical models are not only diffierent in the context of basic assumptions behind the model formulation, rather incomplete or unclear understanding of disease spread for COVID-19 is another burden. In this paper we have proposed and analyzed an ordinary differential equation model to study the COVID-19 disease propagation which consists of fundamentally six compartments. The basic compartments are susceptible, exposed, infected, quarantined, hospitalised and recovered. It is evident that all the exposed individuals can not spread the disease and hence the exposed compartment is divided into two sub-compartments *E*_1_ and *E*_2_ where the individuals belonging to the second compartment can infect the healthy individuals. Once the incubation period is over, exposed individuals are becoming actively infected which means they can infect other individuals. In case of COVID-19 disease, every infected individuals are not developing appropriate symptoms and asymptomatic individuals can recovered from the disease without any serious medical intervention. The infected compartment is divided into two namely asymptomatic infected and symptomatic infected.

There is significant variation in the collection and reporting of infected data and no uniformity is maintained so far due to the rapid spread of the disease. From huge dataset, it is quite difficult to understand when an individual is identified as a COVID-patient but at which category he belongs to (whether an individual belongs to *E*_1_, *E*_2_, *I_a_* or *I_s_* compartment). Without any loss of generality we can assume that the individuals belonging to *I_a_* and *I_s_* are tested to be positive. Once an individual is tested to be positive, he/she needs to follow the appropriate guideline of the country and hence they will be under isolation or quarantine assuming that they do not need any medical intervention through hospitalization. Hence we have assumed that the individuals from *I_a_* compartment directly move to the recovered compartment and hospitalization is required for certain fraction of individuals belonging to sysmptomatic infected and quarantined compartments. COVID-19 related death is assumed for the *I_s_* and *J* compartments only. As the quarantined individuals are monitored on a regular basis and the healthcare service is supposed to be effective, hence every needy individuals can be moved to the hospital as per requirement. Based upon these assumptions we have considered two models here, epidemic model (1) and two group epidemic model (33). Interesting and significant contribution of our work is the consideration of time dependent rate of infection over various periods of time. This variation is adopted into the model in order to capture the effect of lockdown, social distancing etc. which plays a crucial role to reduce the disease spread. The date of implementation of lockdown and the extent of lockdown varies from one country to another. Total lockdown is rarely implemented rather most of the European countries went to step-by-step lockdown and sometimes they imposed complete lockdown at some states and/or provinces instead of total lockdown accross the country.

In this present work we have considered different ranges of days over which the rate of infection varies in order to match the simulation results with the cumulative number of reported infected individuals for four countries. The magnitudes of *β*(*t*) over various intervals are obtained through fitting the simulated result with data but the choice of different periods remain in our hand. One can argue that the obtained results might change with a variant assumption but at the same time we can claim the appropriate nature of varying infectivity is not much clear yet. In order to show the effectiveness of our analysis, the fitting is done for the data from four different countries. It is important to mention here that the fitting with daily data seems to be are not in good agreement (we can see significant variation around the fitted curve) but at the same time we need to keep in mind the irregularity of the reporting protocol. It is a matter of fact that several contries have changed their reporting mechanism time to time alongwith the change in the process of testing. A natural question may arise that we are considering constant *β*(*t*) over different time intervals, so what will be the outcome if we match the simulation result with the daily data or 7 days moving average? Answer is that either we need to change *β*(*t*) in daily basis which is not a good idea for fitting the simulation results of ordinary differential equation models with the data or we can observe a significant disagreement with the cumulative number of reported cases. For numerical simulation of the model (1) we have obtained three different values of *β* and we can calculate the controlled basic reproduction numbers accordingly. For the set of parameter values as mentioned in the three columns of Table 3, we can calculate the controlled reproduction numbers as 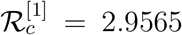, 1.2847, 0.7002. In Section 2 we have calculated the basic reproduction number 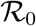, for model (1), apart from the controlled reproduction number 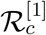. If we calculate the basic reproduction number 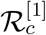 for model (1) with the parameter values given in the first column of Table 3, we find 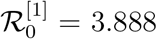 which is quite higher than 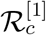. It clearly indicates that the disease can propagate much faster in the absence of the control measures.

The proposed two group epidemic model can capture the situation when one group of people are following the social distancing norm, using face mask appropriately and other group is not obeying the safety norms. We must admit that the data and relevant information for the parameter values involved with the model (33) are not available yet. But the numerical simulation shows that is 10% or more of the population start violating the safety norms then there is a resonable chance that the disease can relapse. It is difficult to predict the size of the next epidemic but our model is capable to capture the realistic scenario. The number of daily reported cases started increasing at some countries like Iran, Spain etc. (see the data avialbale at [3]) compared to the number of reported infections on the days close the end of complete lockdown.

The coefficient of social interaction *K* introduced in Section 4.3 characterizes the intensity of the interaction between susceptible and infectious individuals, and it determines the rate of disease progression. The value *K* = 0 corresponds to complete lockdown, and *K* = 1 to the absence of lockdown. Partial measures of social distancing after the end of lockdown in a number of European countries restrain its value to *K* ≈ 0.1 characterized by a slow growth of the number of infected individuals. Even a slight increase of this coefficient up to *K* = 0.15, which can be expected in September due to the end of vacation season and the beginning of academic year, can lead to an important burst of epidemic.

We have calculated the maximum size of the epidemic for both the models considered in this manuscript and obtained a rough estimate for the endemic steady-state analytically. The maximum size of the epidemic are calculated assuming *β* is constant however the numerical simulations are carried out for non-constant *β* and their variation with time is already explained clearly. Here we justify the analytical findings with supportive numerical results and argue for the effectivity of lockdown and imposition of several social restrictions. We mentioned above that the controlled reproduction number for Germany is 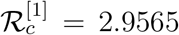 with *β* = *β*_1_ = 3.98. With this value of 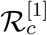, solving the equation (26) we find *x* = 0.06303. With the current population size of Germany which is approximately equal to 82.9 million gives us final size as *S_f_* = 5, 225,187. This result clearly indicates that a huge number of people could be infected if no such restriction was imposed. Similar calculations can be done for other compartments as well for two group model also. But we skip them here for the sake of brevity and will address this issue in our future endevour with appropriate estimates for the values of other parameters involved with the second model.

## Data Availability

The source of the data used in this manuscript are mentioned clearly.

https://www.worldometers.info/coronavirus/

## Acknowledgement

The second author’s (VV) work is supported by the Ministry of Science and Higher Education of the Russian Federation: agreement no. 075-03-2020-223/3 (FSSF-2020-0018).

## Appendix – A

**Proof of Th. 1:** By re-writing the system (1) we have

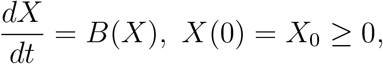

where

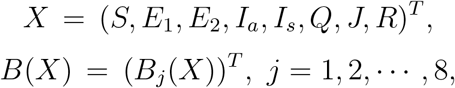

with

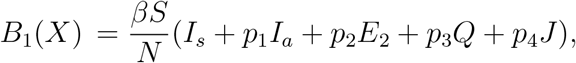

and so on. We note that

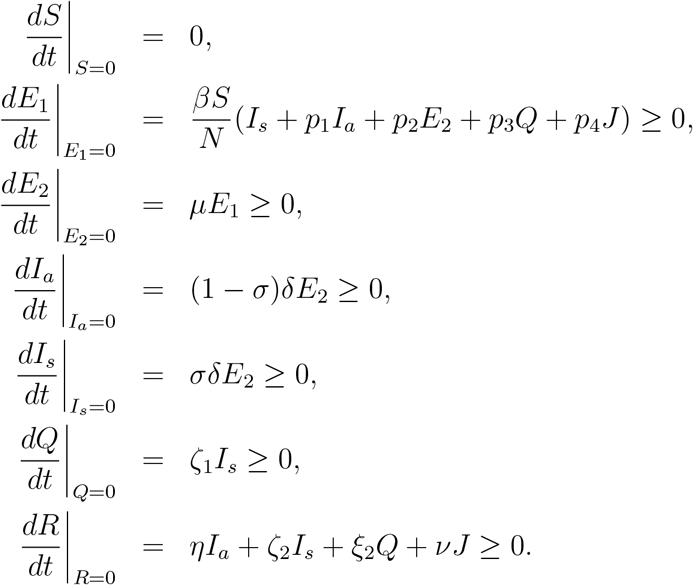

The inequalities mentioned above hold for any point belonging to the interior of 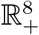 or on the boundary hyper-planes. Solutions starting from a non-negative initial condition and non-negativity of the time derivaties imply that the system (1) is invariant in 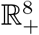.

## Appendix – B

Here we present the numerical simulation results and fitting with the data alongwith the 95% confidence intervals for Germany, Italy, Spain and UK. The data used for these fiiting are available at [3]. The model (1) is used to perform the simulations and fitting with data. Parameterization are also the same as we have explained in Table 1. Details of the parameter values and their estimates for Germany are provided in Table 3. Parameter values and and their estimates for rest of the countries are provided below. It is worthy to mention here that the values of *β*(t) are estimated over requisite number of intervals in order to have a good fit with the cumulative reported number of infected individuals. As a result the number of intervals over which *β*(*t*) is changing its constant magnitude varies from one country to another. In case of Germany, the number of intervals over which *β*(*t*) takes different constant values are three whereas the number of intervals for *β*(*t*) is five when we considered the data for Italy. Also the number days over which the simulations are carried out is not unique as the date implementation of lockdown and its partial withdrawal varies from one country to another. For all the countries, we have started the simulation from the date 15th of February, 2020 and continued approximately up to the end of strict lockdown restrictions.

**Figure 9:**
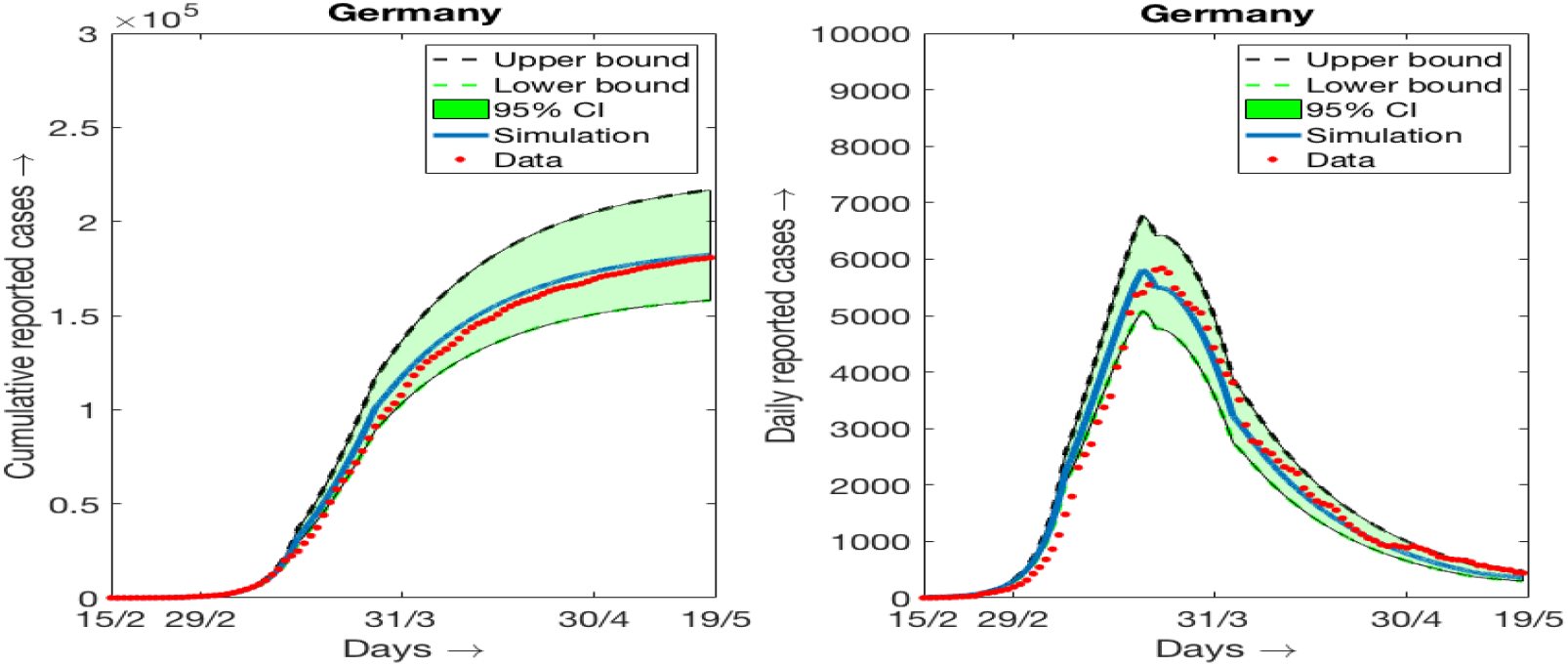
Simulation results and daily reported cases (7 days moving average) and 95% confidence interval for the data of Germany. (a) Cumulative data and simulation results, (b) daily data (7days moving average) and simulation results.

**Table 4:** Best fitted parameter values for Italy.

| Parameter | for 1st 26 days | for next 6 days | for next 7 days | for next 8 days | upto 90th day |
| --- | --- | --- | --- | --- | --- |
| $\beta$ | 4 | 2.89 | 2 | 1.3 | 1 |
| $p_1$ | 0.34 | 0.05 | 0.05 | 0.05 | 0.34 |
| $p_2$ | 0.34 | 0.34 | 0.34 | 0.34 | 0.21 |
| $p_3$ | 0.05 | 0.05 | 0.05 | 0.05 | 0.05 |
| $p_4$ | 0.05 | 0.05 | 0.05 | 0.05 | 0.05 |
| $\delta$ | 1 | 1 | 1 | 1 | 0.96 |
| $\sigma$ | 0.1128 | 0.1 | 0.1 | 0.1 | 0.1 |
| $\eta$ | 0.9 | 0.9 | 0.9 | 0.9 | 0.9 |
| $\rho_1$ | 0.1 | 0.1 | 0.1 | 0.1 | 0.1 |
| $\rho_2$ | 0.07 | 0.07 | 0.07 | 0.07 | 0.07 |
| $\zeta_1$ | 0.07 | 0.07 | 0.07 | 0.07 | 0.07 |
| $\zeta_2$ | 0.1 | 0.1 | 0.1 | 0.1 | 0.1 |
| $\zeta_3$ | 0.14 | 0.14 | 0.14 | 0.14 | 0.14 |
| $\xi_1$ | 0.14 | 0.14 | 0.14 | 0.14 | 0.14 |
| $\xi_2$ | 0.1 | 0.1 | 0.1 | 0.1 | 0.1 |
| $\nu$ | 0.05 | 0.05 | 0.05 | 0.05 | 0.05 |
| $\mu$ | 0.28 | 0.28 | 0.28 | 0.28 | 0.28 |

**Figure 10:**
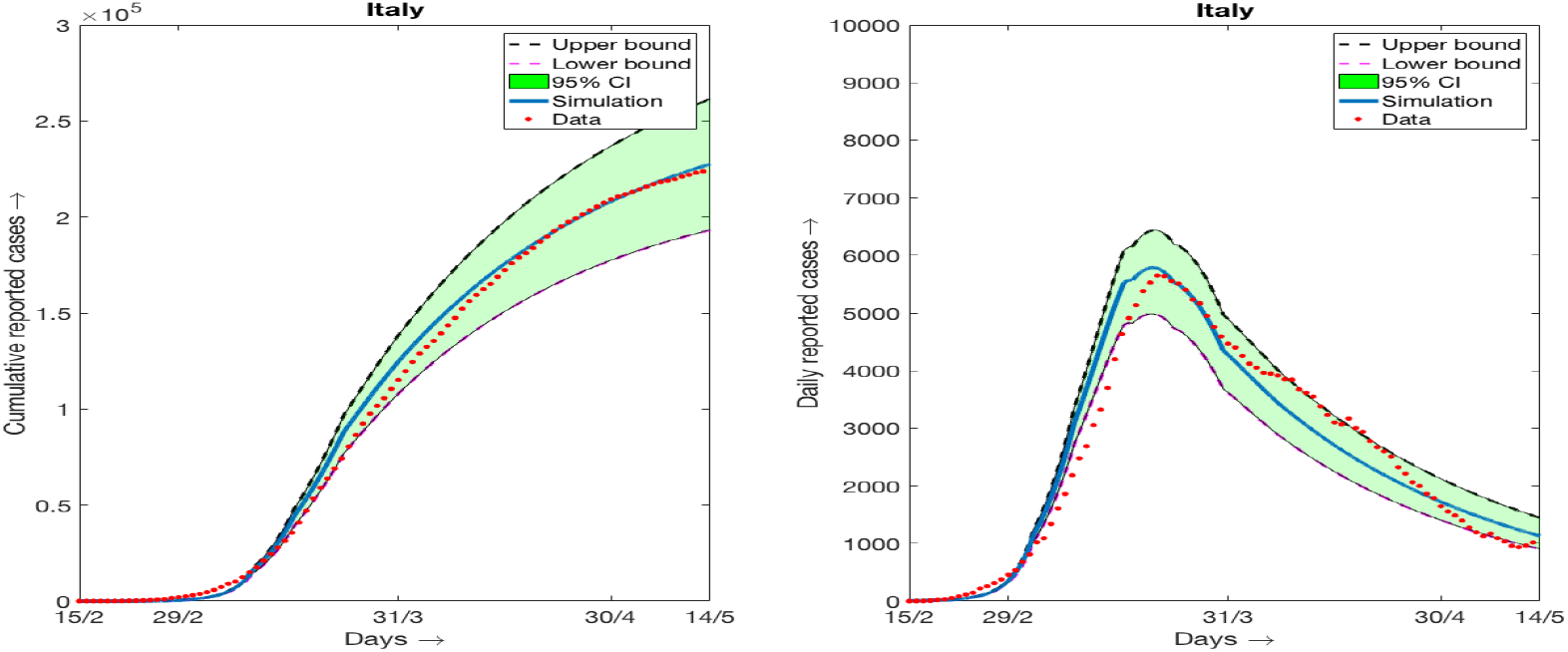
Simulation results and daily reported cases (7 days moving average) and 95% confidence interval for the data of Italy. (a) Cumulative data and simulation results, (b) daily data (7days moving average) and simulation results.

**Table 5:**
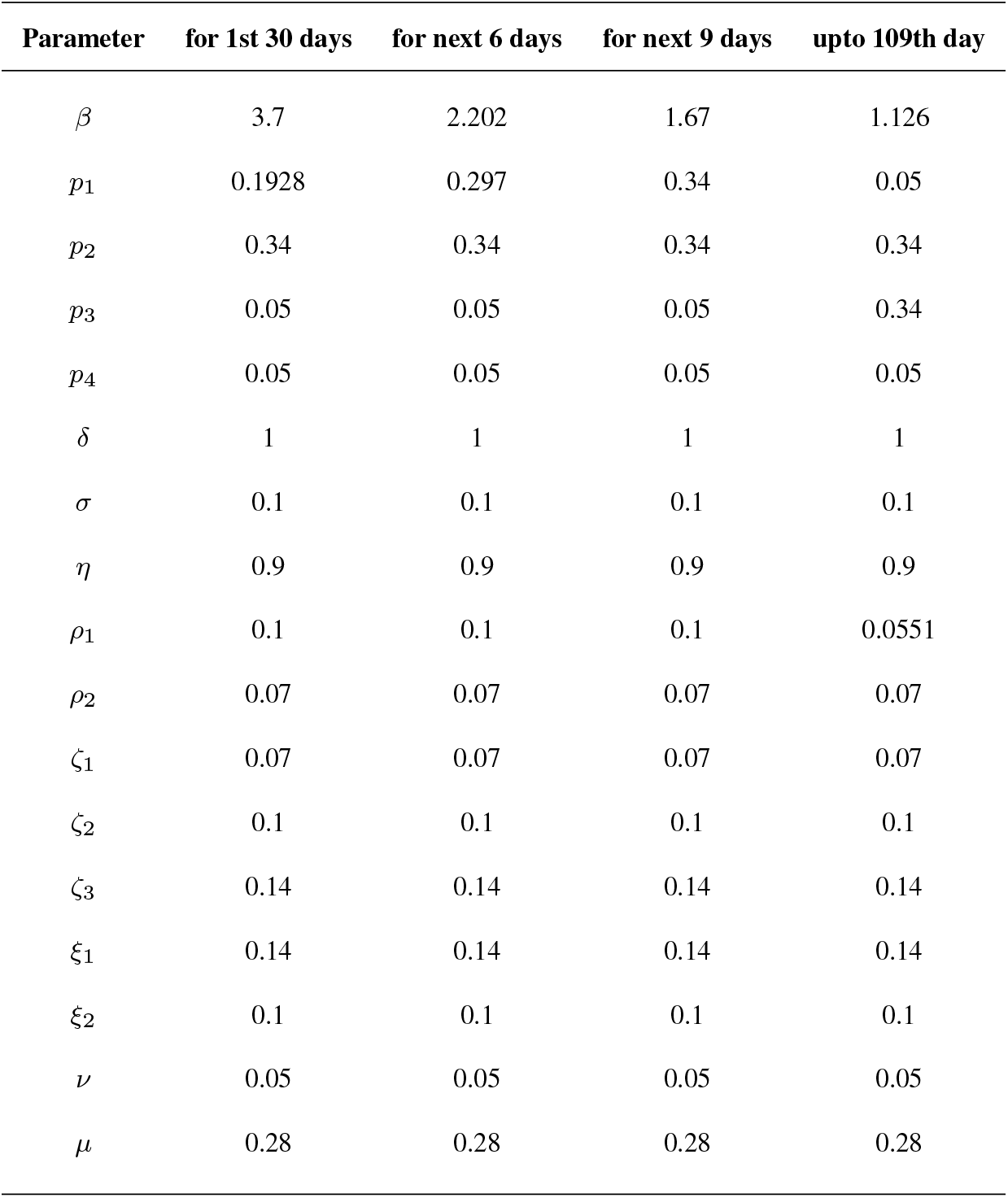
Best fitted parameter values for Spain.

**Figure 11:**
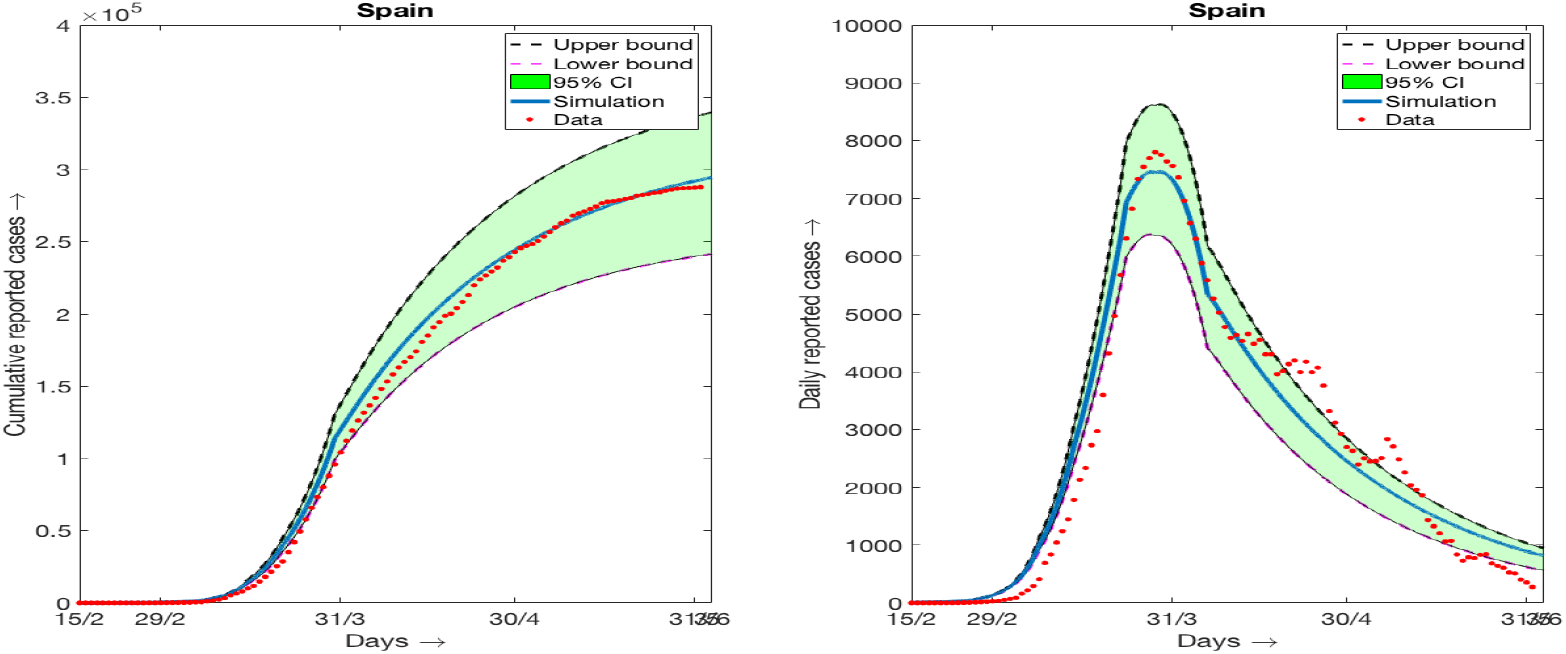
Simulation results and daily reported cases (7 days moving average) and 95% confidence interval for the data of Spain. (a) Cumulative data and simulation results, (b) daily data (7days moving average) and simulation results.

**Table 6:**
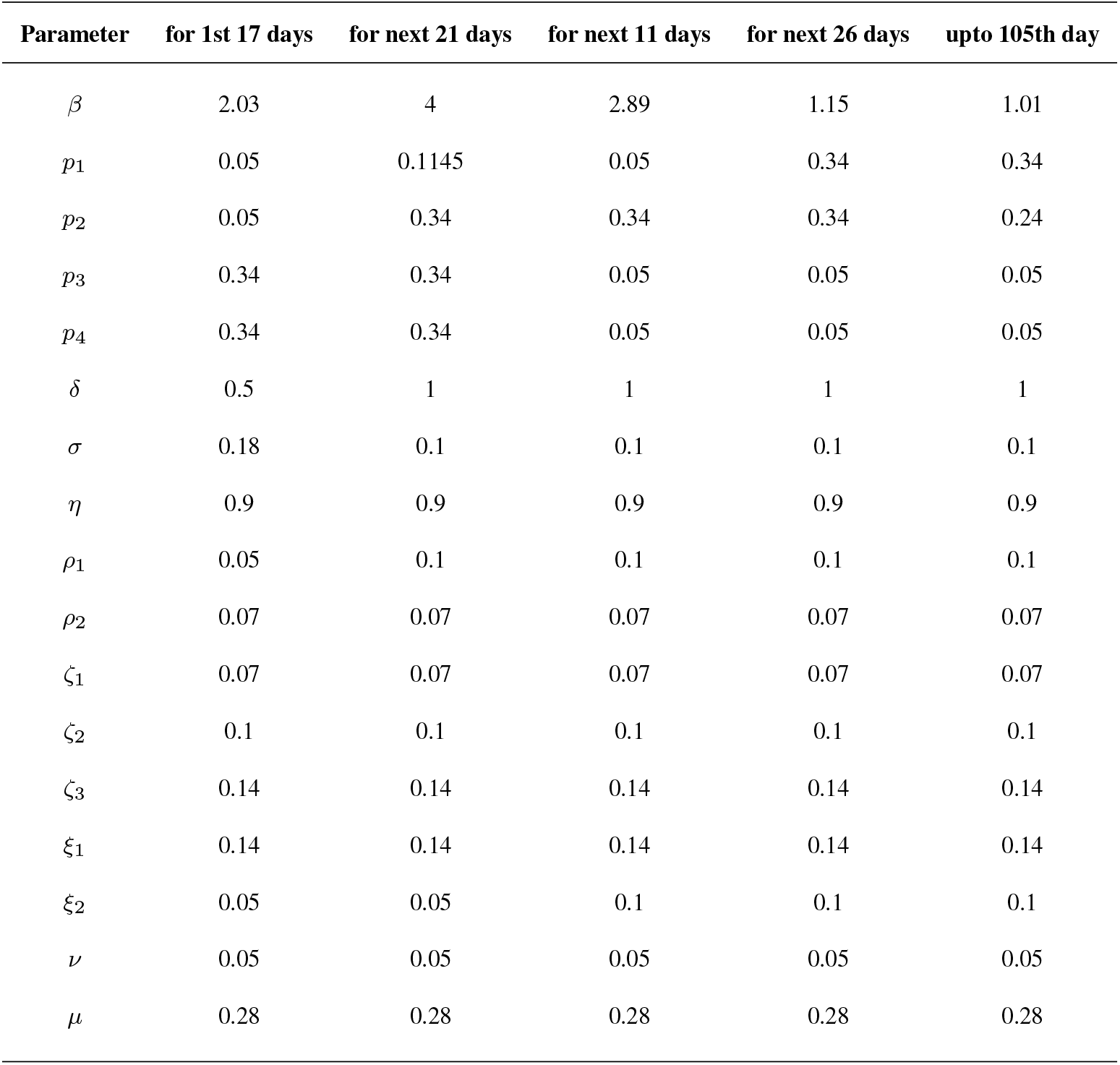
Best fitted parameter values for UK.

| Parameter | for 1st 17 days | for next 21 days | for next 11 days | for next 26 days | upto 105th day |
| --- | --- | --- | --- | --- | --- |
| $\beta$ | 2.03 | 4 | 2.89 | 1.15 | 1.01 |
| $p_1$ | 0.05 | 0.1145 | 0.05 | 0.34 | 0.34 |
| $p_2$ | 0.05 | 0.34 | 0.34 | 0.34 | 0.24 |
| $p_3$ | 0.34 | 0.34 | 0.05 | 0.05 | 0.05 |
| $p_4$ | 0.34 | 0.34 | 0.05 | 0.05 | 0.05 |
| $\delta$ | 0.5 | 1 | 1 | 1 | 1 |
| $\sigma$ | 0.18 | 0.1 | 0.1 | 0.1 | 0.1 |
| $\eta$ | 0.9 | 0.9 | 0.9 | 0.9 | 0.9 |
| $\rho_1$ | 0.05 | 0.1 | 0.1 | 0.1 | 0.1 |
| $\rho_2$ | 0.07 | 0.07 | 0.07 | 0.07 | 0.07 |
| $\zeta_1$ | 0.07 | 0.07 | 0.07 | 0.07 | 0.07 |
| $\zeta_2$ | 0.1 | 0.1 | 0.1 | 0.1 | 0.1 |
| $\zeta_3$ | 0.14 | 0.14 | 0.14 | 0.14 | 0.14 |
| $\xi_1$ | 0.14 | 0.14 | 0.14 | 0.14 | 0.14 |
| $\xi_2$ | 0.05 | 0.05 | 0.1 | 0.1 | 0.1 |
| $\nu$ | 0.05 | 0.05 | 0.05 | 0.05 | 0.05 |
| $\mu$ | 0.28 | 0.28 | 0.28 | 0.28 | 0.28 |

**Figure 12:**
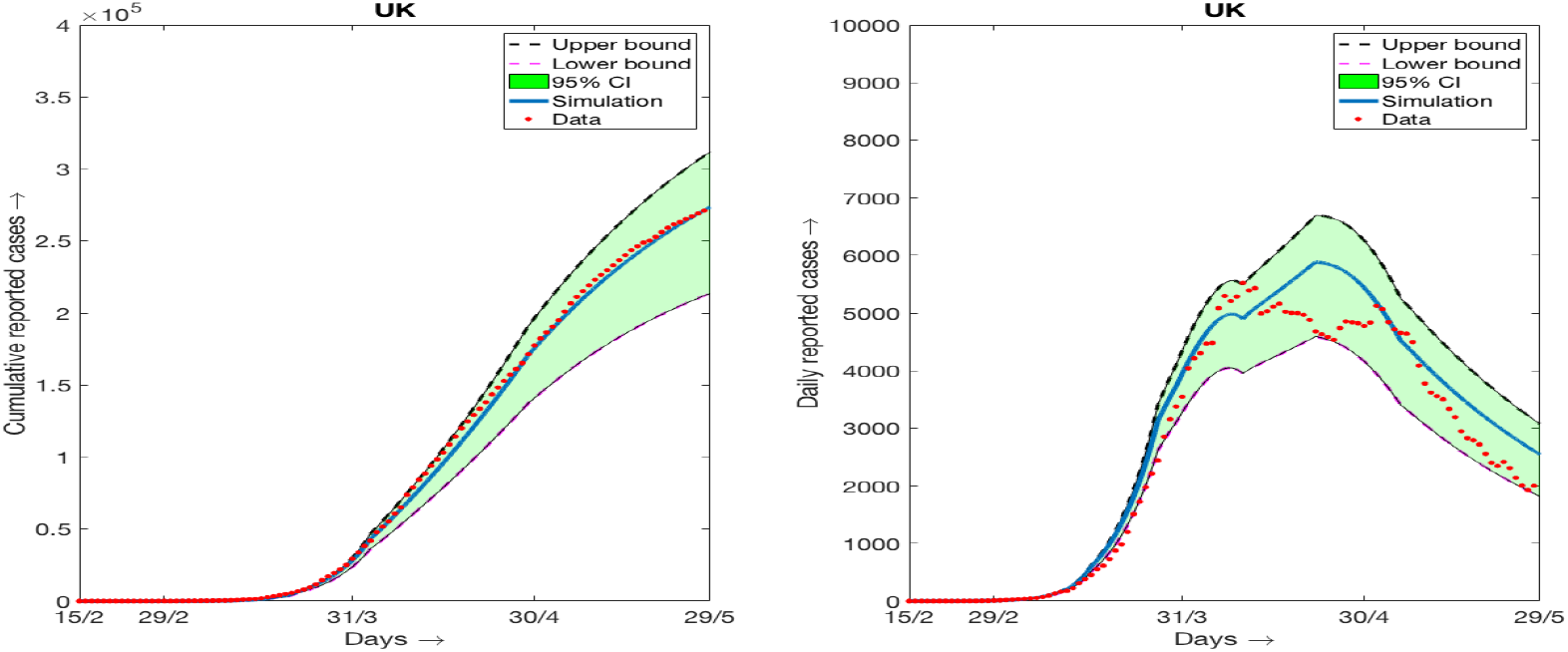
Simulation results and daily reported cases (7 days moving average) and 95% confidence interval for the data of UK. (a) Cumulative data and simulation results, (b) daily data (7days moving average) and simulation results.

## References

[1] World Health Organization.“Coronavirus disease 2019”. cited March 15, 2020. Available: https://www.who.int/health-topics/coronavirus.

[2] Editorial, The continuing 2019-nCoV epidemic threat of novel coronaviruses to global health-The latest 2019 novel coronavirus outbreak in Wuhan, China, Int. J. Infect. Dis., 91 (2020), 264-266.

[3] Worldometer: https://www.worldometers.info/coronavirus

[4] World Health Organization, “Population-based age-stratified seroepidemiological investigation protocol for covid-19 virus infection”, 2020.

[5] N. M. Ferguson et al., “Impact of non-pharmaceutical interventions (npis) to reduce covid-19 mortality and healthcare demand,” London: Imperial College COVID-19 Response Team, 10 (2020), 10.25561/77482.

[6] B. Tang, N. L. Bragazzi, Q. Li, S. Tang, Y. Xiao, J. Wu., An updated estimation of the risk of transmission of the novel coronavirus (2019-ncov). Infect. Dis. Model., 5 (2020), 248-255.

[7] B. J. Quilty, S. Clifford, et al., Effectiveness of airport screening at detecting travellers infected with novel coronavirus (2019-nCoV). Eurosurveillance, 25 (2020), 200080.

[8] M. Shen, Z. Peng, Y. Xiao, L Zhang, Modelling the epidemic trend of the 2019 novel coronavirus outbreak in china, *bioRxiv*, 2020.

[9] A. J. Kucharski, T. W. Russell, C. Diamond, Y. Liu, J. Edmunds, S. Funk, R. M. Eggo, Early dynamics of transmission and control of COVID-19: a mathematical modelling study, Lancet Infect. Dis., 20 (2020), 553–558.

[10] J. Yuan, M. Li, G. Lv, Z. K. Lu, Monitoring transmissibility and mortality of COVID-19 in Europe, Int. J. Infec. Dis., 95 (2020), 311-325.

[11] G. Giordano, F. Blanchini, R. Bruno, P. Colaneri, A. D. Filippo, A. D. Matteo, M. Colaneri, Modelling the COVID-19 epidemic and implementation of population-wide interventions in Italy, Nature Medicine, https://doi.org/10.1038/s41591-020-0883-7.

[12] Q. Lin, S. Zhao, D. Gao, Y. Lou, S. Yang, S. S. Musa, M. H. Wang, Y. Cai, W. Wang, L. Yang, D. He, A conceptual model for the coronavirus disease 2019 (COVID-19) outbreak in Wuhan, China with individual reaction and governmental action, Int. J. Infect. Dis., 93 (2020), 211-216.

[13] T. Chen, J. Rui, Q. Wang, Z. Zhao, J. Cui and L. Yin, A mathematical model for simulating the phase-based transmissibility of a novel coronavirus, Infect. Dis. Poverty, 9 (2020) 1-8.

[14] J. T. Wu, K. Leung, G. M. Leung., Nowcasting and forecasting the potential domestic and international spread of the 2019-ncov outbreak originating in wuhan, china: a modelling study. The Lancet, 395 (2020), 689-697.

[15] B. Tang, X. Wang, Q. Li, N. L. Bragazzi, S. Tang, Y. Xiao, J. Wu., Estimation of the transmission risk of the 2019-nCov and its implication for public health interventions, J. Clin. Med., 9 (2020),462.

[16] J. M. Read, J. R. Bridgen, D. A. Cummings, A. Ho, C. P. Jewell, Novel coronavirus 2019- nCov: early estimation of epidemiological parameters and epidemic predictions. *MedRxiv*, 2020.

[17] S. Zhao, Q. Lin, J. Ran, S. S. Musa, G. Yang, W. Wang, M. H. Wang, Preliminary estimation of the basic reproduction number of novel coronavirus (2019-nCoV) in China, from 2019 to 2020: A data-driven analysis in the early phase of the outbreak. Int. J. Infect. Dis., 92 (2020), 214-217.

[18] J. Chen, Pathogenicity and transmissibility of 2019-nCoV - a quick overview and comparison with other emerging viruses. Microb. infect., (2020), https://doi.org/10.1016/j.micinf.2020.01.004.

[19] R. Singh, R. Adhikari, Age-structured impact of social distancing on the covid-19 epidemic in India. www.arXiv.org, arXiv:2003.12055, 2020.

[20] U, Avila-Ponce de Leon, A. G. C. Perez, E, Avila-Vales, An SEIARD epidemic model for COVID-19 in Mexico: mathematical analysis and state-level forecast, (2020), www.medrxiv.org, https://doi.org/10.1101/2020.05.11.20098517.

[21] J. H. Rojas, M. Paredes, M. Banerjee, Olcay Akman, Anuj Mubayi, Mathematical Modeling & the Transmission Dynamics of SARS-CoV-2 in Cali, Colombia: Implications to a 2020 Outbreak & public health preparedness, (2020), www.medrxiv.org, https://doi.org/10.1101/2020.05.06.20093526.

[22] E. Shim, G. Chowell, Regional variability in time-varying transmission potential of COVID-19 in South Korea, (2020) www.medRxiv.com, https://doi.org/10.1101/2020.07.21.20158923.

[23] A. Srivastava, G. Chowell, Understanding Spatial Heterogeneity of COVID-19 Pandemic Using Shape Analysis of Growth Rate Curves, (2020), www.medRxiv.org, https://doi.org/10.1101/2020.05.25.20112433.

[24] Y. Belgaid, M. Helal, E. Venturino, Analysis of a model for Coronavirus spread, Mathematics, 8(5), (2020), 820.

[25] J. Dolbeault, G. Turinici, Heterogeneous social interactions and the Covid-19 lockdown outcome in a multi-group SEIR model, Math. Model. Nat. Phenom., 15 (2020), 36.

[26] M. Kochanczyk, F. Grabowski, T. Lipniacki, Dinamics of Covid-19 pandemic at constant and time-dependent contact rates, Math. Model. Nat. Phenom., 15 (2020), 28.

[27] S. G. Krantz, P. Polyakov, A. S. R. S. Rao, True epidemic growth construction through harmonic analysis, J. Theor. Biol., 494, (2020), 110243.

[28] S. Sinha, Epidemiological dynamics of the COVID-19 pandemic in India: an interim assessment, Stat. Appl., 18 (2020), 333–350.

[29] H. R. Thieme, Mathematics in Population Biology, Princeton University Press, Princeton, 2003.

[30] A. V. Emmanuelle: Lifting the COVID-19 lockdown: different scenarios for France, Math. Model. Nat. Phenom., (In press) 2020.

[31] L. D. Domenico, G. Pullano, C. E. Sabbatini, P. Y. Boelle, V. Colizza: Expected impact of reopening schools after lockdown on COVID-19 epidemic in Ile-de-France, (2020), www.medrxiv.org, https://doi.org/10.1101/2020.05.08.20095521.

[32] U. Avila-Ponce de Leon, A. G. C. Prez, E. Avila-Vales, An SEIARD epidemic model for COVID-19 in Mexico: mathematical analysis and state-level forecast, (2020), www.medrxiv.org,https://doi.org/10.1101/2020.05.11.20098517.

[33] P. van den Driessche, J. Watmough, Reproduction numbers and sub-threshold endemic equilibria for compartmental models of disease transmission, Math. Biosci., 180 (2002), 29-48.

[34] O. Diekmann, J.A.P. Heesterbeek, J.A.J. Metz, On the definition and the computation of the basic reproduction ratio 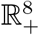 in models for infectious diseases in heterogeneous populations, J. Math. Biol., 28 (1990), 365–382.

[35] F. Brauer, C. Castillo-Chavez, Z. Feng, Mathematical Models in Epidemiology, Springer, New York, 2019.

[36] M. Martcheva, An Introduction to Mathematical Epidemiology, Springer, New York, 2015.

[37] V. Andreasen, The final size of an epidemic and its relation to the basic reproduction number, Bull. Math.Biol., 73 (2011) 2305-2321.

[38] R. J. Freund, W. J. Wilson, D. L. Mohr, Statistical Methods, Elsevier, Canada, 2010.

[39] C. T. Kelley, Iterative Methods for Optimization, SIAM, Philadelphia, USA, 1999.

[40] D. Caccavo, Chinese and Italian COVID-19 outbreaks can be correctly described by a modified SIRD model, (2020) www.medrxiv.org, https://doi.org/10.1101/2020.03.19.20039388.

